# Single-Cell RNA Sequencing of Terminal Ileal Biopsies Identifies Signatures of Crohn’s Disease Pathogenesis

**DOI:** 10.1101/2023.09.06.23295056

**Authors:** Monika Krzak, Tobi Alegbe, D Leland Taylor, Gareth-Rhys Jones, Mennatallah Ghouraba, Michelle Strickland, Bradley T Harris, Reem Satti, Kenneth Arestang, Lucia Ramirez-Navarro, Nilanga Nishad, Kimberly Ai Xian Cheam, Marcus Tutert, Matiss Ozols, Guillaume Noell, Steven Leonard, Moritz J Przybilla, Velislava Petrova, Carla P Jones, Noor Wana, May Xueqi Hu, Jason Skelton, Jasmin Ostermayer, Yong Gu, Wendy Garri, Biljana Brezina, Charry Queen Caballes, Daniele Corridoni, Miles Parkes, Vivek Iyer, Cristina Cotobal Martin, Rebecca E McIntyre, Tim Raine, Carl A Anderson

## Abstract

Crohn’s disease (CD) is a chronic inflammatory bowel disease exhibiting substantial heterogeneity in clinical presentation and response to therapy. To explore its molecular basis, we developed IBDverse, the largest single-cell RNA sequencing (scRNA-seq) dataset of terminal ileal biopsies, profiling over 1.1 million cells from 111 CD patients and 232 healthy controls. This resource integrates discovery and replication cohorts for robust identification of CD-associated cell types, genes, and pathways. We uncovered epithelial changes marked by interferon-driven MHC-I upregulation, persisting in progenitors after macroscopic inflammation resolution. *ITGA4^+^* macrophages were identified as key inflammatory drivers, showing enriched JAK/STAT signaling and cytokine expression (IL-6, IL-12, IL-23). Heritability analysis linked inflammatory monocytes and macrophages to CD susceptibility, implicating resident and recruited immune cells in pathogenesis. These findings establish a comprehensive cellular and molecular framework for CD, offering new insights into disease mechanisms and therapeutic opportunities.

## Introduction

Crohn’s disease (CD) is a debilitating inflammatory bowel disease (IBD) characterised by chronic relapsing and remitting inflammation of the gastrointestinal tract. The biological basis of CD is incompletely understood, although it is hypothesised to be caused by an overactive immune response to commensal gut bacteria in genetically predisposed individuals. Although inflammation is most commonly observed in the terminal ileum, CD exhibits significant heterogeneity in disease location, severity, and behaviour, both between patients and within patients over time. While therapies targeting cytokines such as TNF, IL-12 and IL-23 have improved clinical outcomes for some patients, primary non-response and secondary loss of response to treatment remains high, with 15% of CD patients requiring surgical intervention within five years of diagnosis [1], [2]. Consequently, there is an urgent need to understand better the aetiology of CD in order to broaden therapeutic opportunities.

Genetic association studies have identified over 320 loci associated with IBD susceptibility [3], [4], [5]. Together, these associations have suggested that a broad range of immune, epithelial and stromal cell genes and pathways are involved in CD pathogenesis. However, it is often challenging to draw novel biological insights from these discoveries because most of these disease-associated variants reside in the non-coding genome and it is unclear which genes and pathways they dysregulate. While functional genomic studies of disease-relevant tissues and cell types have shown utility for identifying CD effector genes, pathways and cell types [6], [7], [8], [9], [10], [3], progress has been hampered by their restriction to whole tissues or a limited number of cell types and states.

Single-cell RNA sequencing (scRNA-seq) technologies overcome this limitation by providing a high-throughput means to dissect complex tissues at the resolution of single cells and cell types. ScRNA-seq atlases of the gastrointestinal tract have already made some important discoveries, including the pathogenic remodelling of mesenchymal cells [11] and *CD8^+^* T cells [12] in ulcerative colitis (UC), another common form of IBD, as well as the identification of *BEST4^+^* enterocytes that are crucial in maintaining luminal pH in both UC and CD [13], [14], [15], and identification of metaplastic cells in IBD [16]. Falling scRNAseq costs have begun to enable cell-type resolution comparisons of gene-expression between groups of samples, for example between different gut locations [17], between disease cases and unaffected controls [14], [17] or between responders and non-responders to a drug [18]. While these early discoveries showcase the potential for scRNA-seq-based differential gene-expression studies to deliver biological insights into disease, power to detect differentially expressed genes has been constrained by the small sample sizes used to date. Furthermore, the clinical and biological heterogeneity of IBD, along with the technical noise inherent in scRNA-seq data [19], increases the risk of false-positive associations due to confounding and further limits power to detect true associations. A large scRNA-seq study of gastrointestinal samples ascertained from hundreds of CD patients and healthy controls would thus be a key resource to identify genes, pathways and cell types that are reproducibly associated with CD and related phenotypes.

To address this need, we present IBDverse, a scRNA-seq data resource of over 1,185,000 cells isolated from terminal ileum biopsies from 111 patients with Crohn’s disease and a history of current or previous terminal ileitis and 232 healthy controls. This study, which comprises both a discovery and replication cohort, is uniquely powered to identify gastrointestinal cell types, genes and pathways reproducibly associated with CD. Using these data we identify and replicate genes that are aberrantly expressed in CD, plus those where expression is specific to given cell types and cellular processes. We then identify which of these cell types and processes are likely to play a causal role in disease by quantifying their enrichment within IBD genetic association signals. The terminal ileal cell transcriptional data and associated phenotypic information from the IBDverse are available at zenodo DOI: https://zenodo.org/records/14276773 (Data availability) and the expression atlas of gastrointestinal cell types is accessible at https://www.ibdverse.info/.

## Results

### Large scale single-cell sequencing of terminal ileal biopsies identifies 57 cell clusters comprising epithelial, immune and mesenchymal cell types

Terminal ileal (TI) biopsies were collected from 343 patients during ileo-colonoscopy, comprising 232 biopsies from healthy controls and 111 biopsies from CD patients with either active inflammation in the TI or a documented history thereof (Fig. 1a). Baseline demographics of sex and smoking status were not associated with disease status (p > 0.001, Fisher’s exact test). CD patients were significantly younger than the healthy controls by an average of 8 years (41±12 vs 49±13 years, p<0.001, t-test) (Table 1). The magnitude of TI inflammation was assessed using inflammatory components of the simplified endoscopic score of CD activity (SES-CD) to measure ulceration and mucosal surface involvement. The CD biopsies were subsequently classified according to endoscopic severity of disease using a score based upon three inflammatory components (inflamed surface, ulcer size, ulcerated surface) of the validated “simple endoscopic score for CD” (SES-CD), applied to the terminal ileal segment by a single central reader (TI-SES-CD score). This resulted in 64 inflamed (TI-SES-CD≥3) and 47 uninflamed (TI-SES-CD<3) biopsies from 111 individuals (biopsies are unpaired). Clinical information and metadata for the participants are provided in Table S1.

**Fig. 1.**
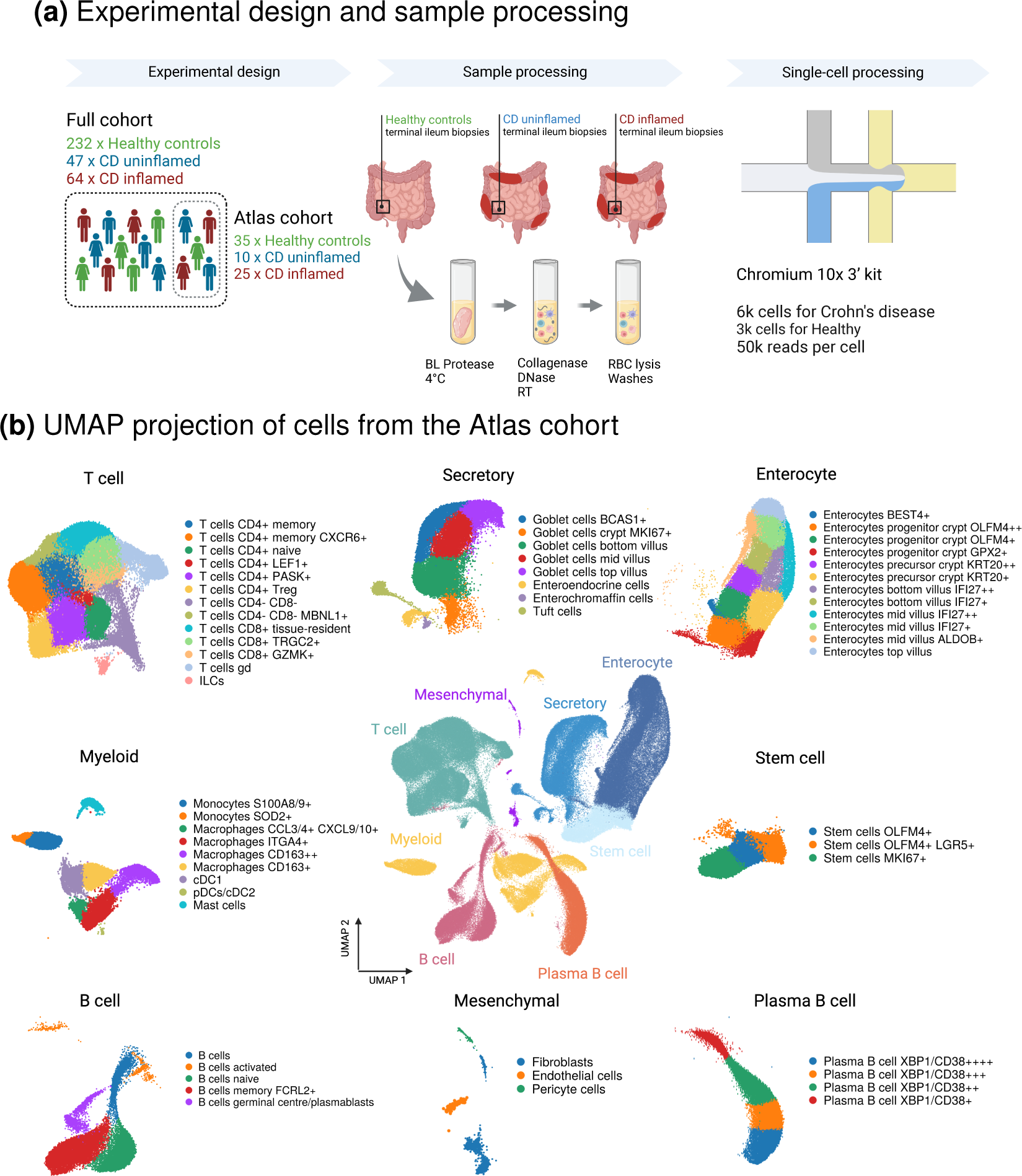
Single-cell expression atlas of the terminal ileum in healthy controls and patients with Crohn’s Disease. **(a)** Terminal ileum biopsies from 343 individuals were dissociated to single cells on ice in Hank’s balanced salt solution (HBSS) containing Bacillus Licheniformis (BL) protease. This was followed by a brief incubation in collagenase and then red blood cell (RBC) lysis buffer. Single cell suspensions were then profiled with the Chromium 10X 3’ kit (Methods). **(b)** Centre: Uniform Manifold Approximation and Projection (UMAP) of *∼*216k cells from the atlas cohort that meet quality control criteria (Methods), with eight colors representing the primary cell populations. Surrounding the central UMAP, these major populations are further subdivided into 57 distinct cell subtypes.

**Table 1.** Demographics of healthy and CD patients in the IBDverse. Absolute number and proportions of demographics across IBDverse. Inflamation was determined based on TI endoscopic score for CD (TI-SES-CD), an aggregated score quantifying the degree of inflammation at several points in the TI. The values of the TI-SES-CD range from 0 to 9 (9 indicating the most severe inflammation). Patients were stratified into two groups: inflamed TI-SES-CD ≥ 3 and uninflamed TI-SES-CD < 3. A significant difference in mean age was observed between healthy controls and CD patients (p < 0.001, t-test). SD=standard deviation.

|  | Healthy (%) | Crohn's Disease (%) | P-value |
| --- | --- | --- | --- |
| N | 232 | 111 |  |
| Inflamed | 0 (0.0) | 64 (57.7) | <0.001 |
| TI-SES-CD |  |  |  |
| [0 – 3) | 232 (100.0) | 47 (42.3) |  |
| [3 – 6) | 0 (0.0) | 50 (45.0) |  |
| [6 – 9) | 0 (0.0) | 14 (12.6) |  |
| Sex = M | 118 (50.9) | 48 (43.2) | 0.205 |
| Smoking Status |  |  | 0.208 |
| No | 178 (76.7) | 77 (69.4) |  |
| Yes | 25 (10.8) | 21 (18.9) |  |
| Ex-Smoker | 25 (10.8) | 12 (10.8) |  |
| Vape Smoker | 4 (1.7) | 1 (0.9) |  |
| Mean Age (SD) | 49.27 (13.31) | 40.87 (12.12) | <0.001 |

All 343 ileal biopsies were processed using a tissue dissociation protocol performed on ice [20] to improve epithelial cell isolation by enhancing cell viability and minimizing cellular stress compared to conventional collagenase digestion at 37°C [21]. A representative cell atlassing cohort consisting of 216,376 high-quality cells (Methods) was derived from 70 randomly selected samples (35 CD {uninflamed/inflamed} and 35 controls), with an average of ∼4,100 post-QC cells from CD biopsies and ∼2,000 post-QC cells from healthy biopsies (Fig. 1b). Marker genes for each of the 57 unique cell clusters were identified using Wilcoxon rank-sum tests, and manual annotations were performed by integrating literature references and lineage-defining gene expression profiles (Fig. S1; Table S2; Methods). These 57 clusters defined three major compartments, comprising eight major cell populations: epithelial cells (enterocytes, secretory, and stem cells), immune cells (T, B, plasma B cells, and myeloid cells), and mesenchymal cells. Positional marker genes from a spatial transcriptomic study of mouse jejunum [22] were employed to annotate enterocytes and secretory cells along the crypt (precursor/progenitor) and villus (bottom/middle/top) axis (Fig. S2). Gene expression signatures of all 57 cell types were then used to build a cell type classifier to annotate cells from the remaining 273 samples not included in the atlasing cohort.

As expected, the cellular composition was altered in CD, with overrepresentation of cell types known to accumulate in intestinal inflammation. For example, *S100A8/9*^+^ monocytes and *CXCL9/CXCL10*^+^ macrophages were effectively absent (<2%) in health (Fig. S3). In disease, these cell-types comprised 25% and 7%, respectively, of all cells in the myeloid compartment, in keeping with their known role in pathogen/inflammatory response [18], [23], [24], [25]. Similarly, four clusters of enterocytes (*GPX2^+^* progenitor crypt, *KRT20^+^* precursor crypt, *IFI27*^++^ mid villus and *IFI27^+^*^+^ bottom villus) were all almost exclusively found in CD (Fig. S3), and displayed transcriptional changes associated with barrier-response to injury. For example, both the “*IFI27*^++^ bottom and middle villus” clusters were denoted by high expression of *CEACAM20*, a sensor of gram negative bacteria and driver of IL-8 release through SAP-1 phosphorylation, antimicrobial peptides (*REG1/3B*) and cytokine responsive elements (*NOS2* and *HSD3B2*), though expression of these markers was consistently lower in the middle versus bottom villus cluster. *HSD3B2*, a steroid synthetase essential for progesterone production, has been shown to modulate local cytokine and repair mechanisms, highlighting the epithelium as not simply a physical barrier, but a key component of the inflammatory cascade, including extra-adrenal steroid production [26], [27].

The small sample sizes commonly used in single-cell differential gene expression studies, combined with the technical and biological variability inherent in single-cell analyses of complex tissues, have made it difficult to validate discoveries of differentially expressed genes and pathways. This has led to claims that the field of single-cell sequencing is facing a reproducibility crisis [28], [29], [30]. To directly assess the replicability of our analyses, all 343 samples in the study were randomly allocated to either a ‘discovery’ or ‘replication’ cohort. First, the 70 samples in the atlasing cohort were equally distributed at random between the two cohorts to minimize bias introduced by variation in the accuracy of cell atlasing. The remaining samples not included in the atlasing cohort were then randomly assigned to either cohort. Ultimately, the discovery cohort consisted of 57 CD (29 with inflamed and 28 with uninflamed CD) and 114 healthy individuals, while the replication cohort included 54 CD (35 with inflamed and 19 with uninflamed CD) and 118 healthy controls (Fig. 2a). No significant differences were observed in baseline demographics between the two cohorts (Table S3). The cell-type classifier was then applied to auto-annotate all 611,992 QC passing cells in the discovery cohort and 573,869 cells in the replication cohort (Methods).

**Fig. 2.**
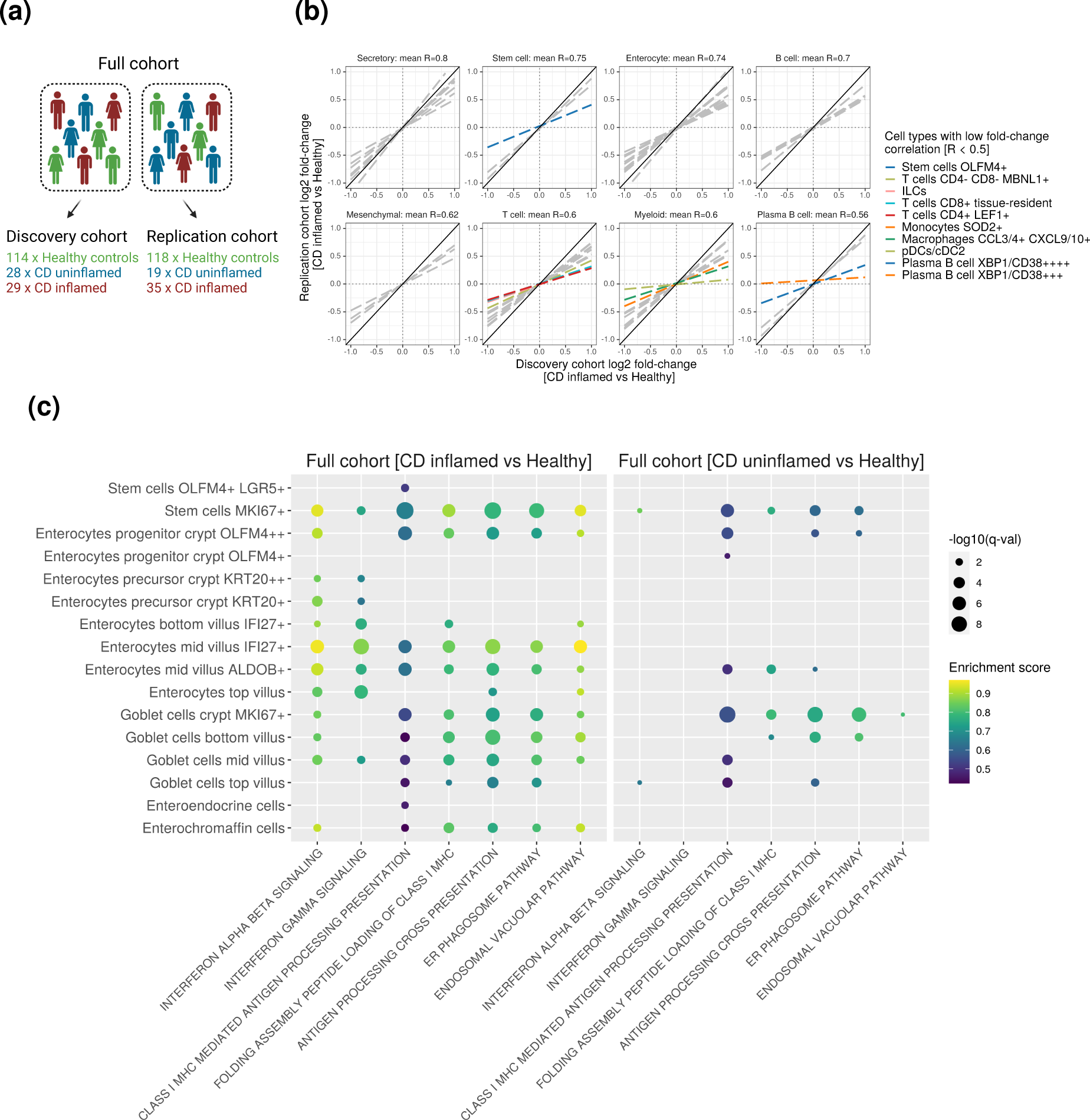
Replicable differential gene expression signatures in CD epithelial cells enriched for interferon signaling and MHC I antigen presentation. **(a)** Design of discovery and replication cohorts; i) first, 70 samples in the atlasing cohort were equally distributed at random between discovery and replication cohorts ii) the remaining 273 samples not included in the atlasing cohort were then randomly assigned to either cohort **(b)** Linear regression (dashed lines) between log2 fold changes of differentilly expressed genes (without thresholding, as outlined in Methods) in the discovery (x-axis) and replication (y-axis) datasets. The reported mean R represents the average of regression coefficients calculated across cell types within each major cell population. Highlighted cell types with low fold-change correlation (R < 0.5, Table S5) **(c)** Gene set enrichment analysis was performed on differential gene expression z-scores derived from the full cohort, comparing CD inflamed samples (n=64) versus controls (n=232) and CD uninflamed samples (n=47) versus controls (n=232). Results for epithelial cell types with high fold-change correlation (R *≥* 0.5).

Within the atlasing cohort we had both the original and auto-annotation cell labels, and were able to quantify the agreement. Overall, the auto-annotation classifier performed well, with similar agreement in annotation between discovery and replication cohorts, demonstrated by high correlation in specifically expressed genes between the original and auto-annotated clusters (Pearson’s R > 0.77) (Fig. S4; Methods). As expected, the accuracy of the autoannotator varied between cell-types, with 49 cell types showing an annotation concordance of more than 50% (Fig. S5). Eight cell types exhibited poor annotation concordance (with fewer than 50% of cells concordantly annotated) - plasmacytoid dendritic cells and conventional dendritic cells type 2 (pDCs/cDC2), *GZMK*^+^ and tissue-resident *CD8*^+^ T cells, *OLFM4*^+^ stem cells, *XBP1/CD38*^+++^ and *XBP1/CD38*^++++^ plasma B cells, and top / mid villus *IFI27*^+^ enterocytes. Somewhat reassuringly, misannotation was often between transcriptomically similar cells. For example, misannotated plasma B *XBP1/CD38*^+++/+^ cells were almost exclusively classified as plasma B *XBP1/CD38*^++^ cells, and misannotated *OLFM4^+^*stem cells were predominantly classified as *OLFM4^+^* epithelial progenitors.

### Identification and replication of dysregulated genes in Crohn’s disease

Across each of the 57 clusters, we compared mean gene expression between inflamed CD biopsies and uninflamed biopsies from healthy individuals. To directly assess replicability, this was performed within both the discovery and replication cohorts independently, and across the two cohorts combined (full cohort) (Fig. 2a). We found 4241 unique differentially expressed genes (DEGs) in the discovery cohort, 4166 unique DEGs in the replication cohort, and 5385 unique DEGs in the full cohort (Table S4; false discovery rate [FDR] < 5%). As expected, the total number of DEGs for each major cell population was positively correlated (R_Discovery_ = 0.66, R_Replication_=0.52, R_Full_=0.73) with the number of sequenced cells (Fig. S6a), albeit with variation at the level of individual cell types (Fig. S6b). For example, secretory epithelial cells, enterocytes and T cells had the highest number of DEGs in the full cohort (totals of 6527, 6324 and 4866, respectively), while plasma B cells had only 719 DEGs, despite the large number of sequenced cells (∼71k in full cohort).

We hypothesised that power to detect and replicate DEGs across our cohorts might be affected by the transcriptional variability (either technical or biological) of individual cell types. To assess this, we compared the gene-expression fold change estimates from our differential gene expression tests between our discovery and replication cohorts (Fig. 2b). When comparing the major cell populations, epithelial cells were the most replicable (mean R=0.76) and plasma B cells the least (mean R=0.56). However, while eight of the top ten most replicable cell types were within the epithelial cell lineage, all major cell populations showed wide variation in fold-change replicability between constituent cell-types. For example, the fold-changes estimates were replicable for gamma-delta T cells (R=0.82), but less so for *LEF1^+^* T cells (R=0.28) or *CD8^+^* tissue-resident T cells (R=0.32) (Table S5). There was also significant variation in fold-change correlation between individual cell-types in the myeloid compartment, with *ITGA4^+^*macrophages demonstrating the best agreement (R=0.88) and pDCs/cDC2s the least (R=0.12). Four of the ten cell types with a fold-change correlation less than 0.5 (pDCs/cDC2, tissue-resident *CD8*^+^ T cells, *OLFM4^+^*stem cells, *XBP1/CD38^+++^* and *XBP1/CD38^++++^* plasma B cells) showed poor annotation consistency between the manual and auto-annotation approaches in the atlasing cohort (fewer than 50% of cells concordantly annotated). This highlights the importance of accurate and consistent cell annotation for single-cell RNA-seq studies to ensure reliable differential gene expression analysis. Two myeloid cell types (*SOD2^+^*monocytes and *CCL3/4^+^ CXCL9/10^+^* macrophages) showed poor fold-change correlations (R<0.5) despite high consistency between the manual and auto-annotation (>98% of cells concordantly annotated). The poor replicability observed for these cell-types was instead likely underpinned by imbalance in the number of cells from cases and controls. Together, these factors make it difficult to accurately estimate mean gene-expression for these cell-types, particularly in the controls, hindering power to detect differences in mean expression correlated with disease status.

Ultimately, only 44% of DEGs (FDR < 5%) detected in the discovery cohort were also significantly dysregulated (FDR <5%) in the same cell type, with the same direction of effect, in the replication cohort. As anticipated, the correlation in gene-expression log-fold changes in our discovery and replication cohort underpinned the replication rate of differentially expressed genes. Consistent with this, the replicability of a given gene is highly cell-type dependent, with epithelial cells, particularly precursors/progenitors and stem cells demonstrating the most consistent differentially-expressed genes between cohorts.

### The Ileum undergoes both pan-epithelial and cell-type specific changes in CD inflammation

To maximise power to identify biological pathways dysregulated in disease, we undertook gene set enrichment analysis (GSEA) across the differential gene expression results from our full cohort (Methods). To limit false-positive enrichments, we focussed this analysis on the 47 cell types with greatest replicability in our differential gene expression (DEG) analysis (Fig. 2b; Table S5; threshold R ≥ 0.5). Epithelial cells demonstrated the strongest correlation in fold change estimates (R=0.76) of the three major cellular compartments, and in the full cohort we observed both cell-type specific and pan-epithelial pathway enrichments (Fig. 2c).

The most widespread pan-epithelial signature in CD inflamed versus healthy control samples was upregulation of “IFNα/β signalling”, detected in twelve epithelial cell types along the entire crypt-villus axis (Fig. 2c). Leading edge analysis showed that the genes driving the observed enrichments for “IFNα/β signalling”, namely *IFI27, HLA-A/B/C/E/F, IFITM3, PSMB8, STAT1* and *IRF1,* were differentially expressed across all twelve cell types (Table S6). Many of these leading edge genes also featured in other significantly enriched pan-epithelial pathways, including *HLA-A/B/C/E/F* and *PSMB8* in the upregulation of “Antigen processing cross-presentation”, “Folding-assembly, peptide loading of class I MHC”, “ER phagosome” and “Endosomal vacuolar pathway”. To determine whether IFNα/β signalling enrichment persisted in patients who had successfully resolved macroscopic inflammation, we undertook differential gene expression analyses across the twelve IFNα/β-enriched cell types, comparing cells from uninflamed CD biopsies to those from healthy controls (Fig. 2c). We did not observe an enrichment of differentially expressed genes in the IFNα/β pathway, except for a weak but statistically significant enrichment in “stem cells *MKI67*” in the uninflamed CD versus healthy biopsy comparison. This suggests that the enrichment of Type I IFN signalling across epithelial cell types in active CD is a widespread barrier response to injury that largely subsides after repair.

Type I interferon (IFN-α/β) and type II interferon (IFN-γ) have long been implicated in the upregulation of MHC-I expression and components of its pathway [31], [32], [33]. Consistent with this, genes differentially expressed between cells from CD inflamed versus healthy biopsies were enriched in the “folding assembly peptide loading of class I MHC” pathway, including (*CANX, CALR, TAPBP, B2M*), across 12 individual epithelial cell types representing all major subtypes (Table S6). However Type II IFN responses, through the “IFNγ signalling” pathway, appeared more restricted, with enrichment preferentially to enterocytes along the entire crypt-villus axis, but not in other epithelial cell types, e.g. secretory cells (Fig. 2c). Given that multiple MHC-I pathways and associated genes were consistently enriched across epithelial cell types, we speculated whether inflammation severity was associated with the strength of this effect. To test this, we fit a differential expression (DE) model for CD patients only stratified according to endoscopic severity of disease using the TI-SES-CD score. In this analysis, “MHC-I mediated antigen processing presentation”, “folding assembly peptide loading MHC-I” and “antigen processing cross-presentation” pathways remained enriched in all epithelial cell types except goblet cell top-villus and enteroendocrine subsets (Fig. S7a). This suggests a widespread dose-dependent effect of inflammation on MHC-I upregulation. Importantly, analyses comparing non-inflamed CD to those from healthy controls showed that this upregulation of MHC-I antigen-presentation persisted even after the resolution of inflammation (Fig. 2c).

Whilst MHC-I is expressed by all cells, MHC-II expression is thought to be restricted to professional antigen presenting cells to limit activation of the adaptive immune system through T cell receptor engagement. However, in the context of inflammation, intestinal epithelial cells have been shown to upregulate MHC-II molecules [18], [13], [34], [35]. Consistent with this, we also observed upregulation of some MHC-II signaling genes (*HLA-DRA, HLA-DRB1, HLA-DPA1, HLA-DRB5, HLA-DQB1* and *HLA-DMB*) but only within specific enterocyte and stem cell subsets (mid villus *ALDOB*^+^, mid villus *IFI27*^+^, progenitor crypt *OLMF4*^++^ and *MKI67*^+^ stem) (Fig. S7b). Thus MHC-II signalling upregulation was less widespread across epithelial cells compared to MHC-I.

Collectively these results suggest widespread changes across epithelial sub-types relating to Type I IFN and MHC-I signalling, with these changes associated with inflammation severity. Importantly, these perturbations in MHC-I signalling may persist in key progenitor cells long after the initial inflammatory stimuli have been removed. Alongside these changes in immune potential in epithelial cells, we also detected evidence of shifts in metabolic function with evidence of alterations in pathways and genes associated with both oxidative phosphorylation and glycolysis (Fig. S7c).

### *ITGA4*^+^ macrophages are enriched for JAK/STAT signalling in ileal CD inflammation

Genetic variants associated with increased expression of *ITGA4* (CD49d) in stimulated monocytes have been associated with increased risk of IBD [3]. *ITGA4* is known to be expressed by a range of circulating lymphocytes, including classical monocytes, where it forms the α4β7 integrin that is the target of the IBD therapeutic, vedolizumab. Anti-α4β7 therapy abrogates blood monocyte MADCAM-endothelium interactions, with circulating levels of α4β7^+^ classical monocytes higher, and tissue monocytes lower, in vedolizumab non-responders [36], [37]. In our terminal ileum cell atlas, *ITGA4* was most highly expressed by two myeloid populations, *ITGA4^+^* macrophages and *CD163^++^*macrophages (Fig. S1).

In patients with active CD, *ITGA4^+^* macrophages constituted a higher mean proportion of myeloid cells within inflamed tissue (16%) compared to healthy controls (10%) (Fig. S3a). These cells also demonstrated the highest reproducibility in our differential gene expression analysis, with a correlation coefficient of R=0.88 (Table S5). Gene set enrichment analysis revealed that differentially expressed genes in *ITGA4^+^* macrophages were enriched across a wide array of cytokine pathways, including interleukins (IL) 1, 4, 6, 9, 10, 12, 13, 20, 21, 23, 27 and 35 (Fig. S3b). These cytokines suggest a predominant role of signalling through receptors of the IL-6 and interferon (IFN) superfamilies, predominantly mediated by Janus kinases (JAK) JAK1/2 and TYK2 [38], [39]. Concordantly, *JAK2* and *STAT1* were the most overexpressed genes in our differential gene expression analysis, with *JAK2* and multiple *STAT* isoforms (*STAT1/2/3*) identified as key mediators in the leading-edge genes across the enriched pathways. Negative regulators of JAK/STAT signalling, such as *PTPN1*/*2,* were also upregulated in *ITGA4^+^* macrophages and were leading edge genes in a number of enriched pathways, including “IL-1 signalling”, “Signalling by interleukins” and “Cytokine signalling in immune system”. Furthermore, we found that the expression of genes in many of these enriched cytokine signalling pathways, including IL-10, IL-12 and IL-20, were positively correlated with inflammation severity (TI-SES-CD score) in *ITGA4^+^* macrophages (Fig. S3b).

Building on the observed correlations between IL-10 and IL-20 signalling in *ITGA4^+^* macrophages and the severity of inflammation in active CD, we further investigated the potential involvement of IFN super-family receptors. Notably, *ITGA4^+^* macrophages upregulated genes associated with proteasome (e.g., PSMA4/5) and immunoproteasome (e.g., PSMB8/9) complex formation, that are known to be dependent on IFN signalling (Fig. 3c). While the proteasome primarily facilitates the normal degradation of proteins, the immunoproteasome is upregulated in response to pro-inflammatory signals such as Type II IFN, enhancing the generation of peptides from pathogen-derived proteins for MHC-I-mediated presentation to *CD8*^+^ T cells. This extensive engagement in protein catabolism and antigen presentation suggests that *ITGA4^+^* macrophages may play a crucial role in amplifying adaptive immune responses in the local tissue environment.

**Fig. 3.**
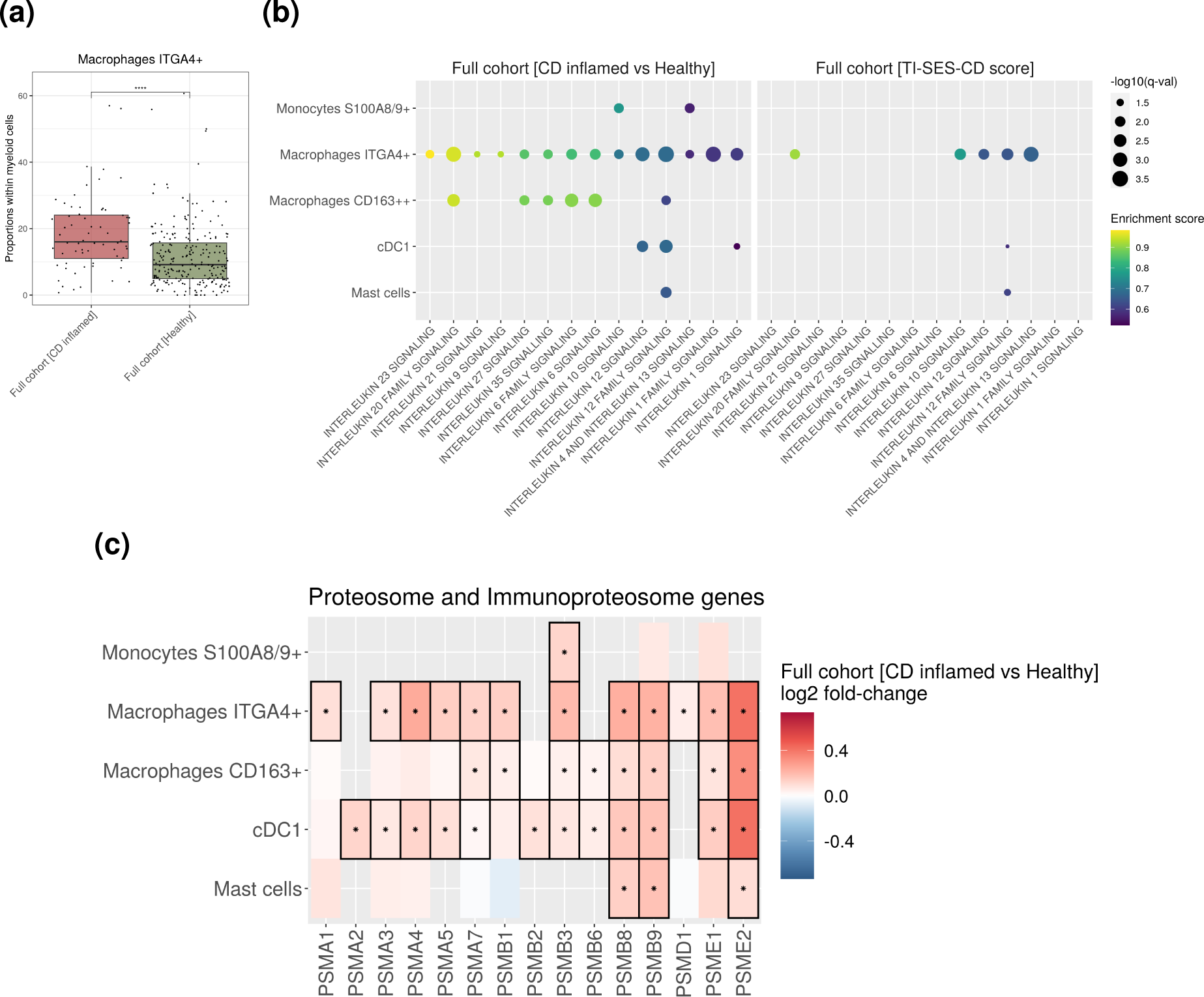
*ITGA4*-positive macrophages upregulate cytokine and proteasome genes during inflammation. **(a)** Proportions of *ITGA4*-positive macrophages within myeloid cells across inflamed CD (n=64) and healthy samples (n=232) (adjusted p values **** < 0.0001, t-test). **(b)** Gene set enrichment analysis of differential gene expression between inflamed CD (n=64) and healthy samples (n=232) and across CD samples with varying severity of inflammation (n=111) - CD samples were stratified based on the simple endoscopic score applied to the terminal ileal segment (TI-SES-CD), with scores ranging from 0 to 9. Enrichment shown for replicable myeloid cell types (R *≥* 0.5, Table S5) **(c)** Fold change and significance of proteasome and immunoproteasome genes differentially expressed in the replicable myeloid cell populations.

Taken together, our findings show that *ITGA4^+^* macrophages are over-represented in CD inflammation and preferentially express an array of cytokine signalling pathways. The specific repertoire of interleukins suggest IL-6 and IFN super-family receptor signalling with resultant JAK activation, particularly JAK2, with inflammation-dependent increases in *IL-10*, *IL-12* and *IL-20*. These cells further demonstrate the effects of Type II IFN signalling, with enrichment for many pathways involved in immunoproteasome-MHC-I communication, suggesting an active role in cytokine sensing, signalling and adaptive immune-cell cross-talk.

### IBD-associated genes are expressed across all major cell populations of the gut

Genome-wide association studies have identified more than 300 regions of the human genome associated with IBD susceptibility. Translating these genetic discoveries into biological understanding and therapeutic hypotheses requires elucidation of the cell types in which they are operative. High resolution single-cell atlases are beginning to deliver these important insights, with previous studies identifying cell types that highly express some IBD-associated genes [17], [40]. To build on these efforts, we compared gene-expression specificity scores across the 57 cell-types in our atlas for 45 high confidence candidate effector genes (Methods) from IBD genome-wide association studies (Fig. 4). All eight major cell populations showed specific expression of at least one IBD effector gene, highlighting the complex cellular architecture of IBD and the absence of a single, dominant cell-type or pathway underlying pathology. For example, *FUT2* was specifically expressed by *IFI27/KRT20*-positive enterocytes (mean specificity = 0.7) but not differentially so between CD cases and controls (Table S4)*. FUT2* co-ordinates synthesis of the carbohydrate HBGA, responsible for ABO blood group system, but these antigens are also secreted in mucosal tissue sites. Indeed, *FUT2* mutations that increase IBD risk have been associated with ‘non-secretor’ status of these antigens, which in turn impair attachment of *Bifidobacteria* and *Lactobacilli*. Abrogation of *FUT2* expression, either via transgenic approaches or naturally occurring genetic polymorphisms, results in dysbiosis and increased susceptibility to inflammation [41], [42]. We show that *FUT2* is predominantly expressed by enterocytes and stem cells (specificity > 0.5) compared to other major cell populations (specificity < 0.36), with expression greatest in precursors/progenitors and falls progressively as enterocytes mature up the crypt.

**Fig. 4.**
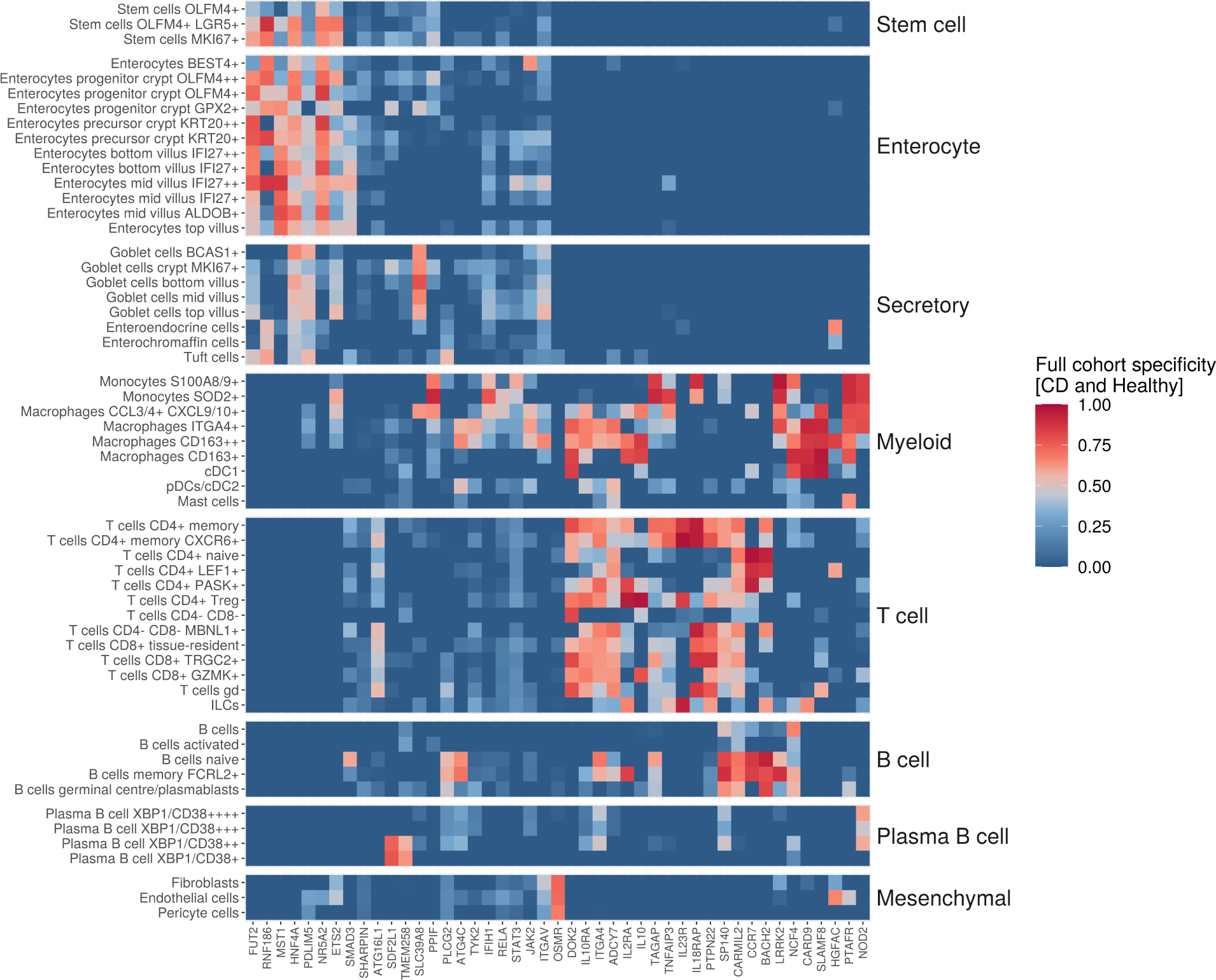
IBD risk genes show lineage-specific expression across terminal ileum cell types. Specificity of IBD risk gene expression across 57 terminal ileum cell-types in the full cohort (n=343). For prioritisation criteria, see Methods.

A small subset of coding mutations in *NOD2* comprise the strongest genetic effects on ileal CD in Western populations, and have been suggested to impair handling of intra-cellular bacteria by myeloid cells [43], [44], [45]. We observed specific expression of *NOD2* in monocytes (*S100A8/9^+^* and *SOD2^+^*) and immature (e.g. *ITGA4^+^* and *CXCL9/10^+^* cells) (mean specificity=0.8), but not resident (e.g. *CD163^+^* and *CD163^++^* cells) macrophages (mean specificity=0.2). Across all of these cell types we found no evidence of *NOD2* differential expression between CD-inflamed biopsies and those from healthy individuals (Table S4). Given that these *NOD2*^+^ cell types are also known to be rapidly recruited from blood as a critical component of the response to intestinal damage [46], impaired bacterial handling in the initial emergency response by these cells may lead to microbial persistence and represent an early triggering event for chronic immune activation. Three of these *NOD2*^+^ cell types (*SOD2*^+^ monocytes, *S100A8/9*^+^ monocytes and *CXCL9/10*^+^ macrophages) were also the sole source of oncostatin M (*OSM*) in our data (mean specificity=0.9). Oncostatin M has been suggested as a potential biomarker and therapeutic target in IBD, that induces an inflammatory programme including *IL6*, *CXCL9* and *CXCL10* in stromal cells, with high pretreatment levels associated with anti-TNF failure [47]. The cognate receptor, *OSMR,* has been shown to be expressed on CD45^−^EpCAM^−^CD31^−^ cells and its expression is co-linear with *COL1A1*, *FAP* and *PDPN* fibroblast markers [47], at whole tissue level. In our study, we confirm cell-specific expressions of *OSMR* within fibroblast, endothelial and pericyte subpopulations of mesenchymal cells (specificity=0.74, 0.72, 0.67, respectively). These observations collectively demonstrate the utility of continuing to build comprehensive cellular atlases to enhance the interpretation of genetic association studies of IBD.

### Heritability enrichment analysis identifies disease relevant cell types for CD and UC

As well as assigning putative effector cell types to specific genetic risk loci, we also sought to identify cell-types of pathologic relevance based upon correlation of cell type specific gene-expression scores with genome-wide maps of genetic susceptibility for CD, UC and IBD. Genome-wide summary statistics for height and educational attainment were used as negative controls in these heritability enrichment analyses [7], [48], [49], [50].

Only cell types of the innate and adaptive immune system showed enrichment of heritability for CD, UC and IBD. Among innate immune cells, specifically expressed genes within *S100A8/9^+^* and *SOD2^+^*monocytes, *CXCL9/CXCL10^+^* macrophages, *CD163^+/++^*and cDC1s were significantly enriched for CD but not UC heritability (Fig. 5; FWER < 0.05). This suggests a potential causal role for cell types involved in both inflammatory responses (*S100A8/9^+^* monocytes, *CXCL9/10^+^*macrophages) and immune tolerance (*CD163^+^* macrophages and cDC1s) in the pathogenesis of CD. Furthermore, myeloid populations with genes significantly enriched for CD heritability, including *S100A8/9^+^* monocytes, *SOD2^+^*monocytes and *CCL3/4^+^ CXCL9/10^+^* macrophages, were significantly reduced in healthy samples (Fig. S8; p adj < 0.05, t-test). Within the adaptive immune compartment, genes specifically expressed by *CD4^+^* memory/*CXCR6^+^* T cells, Tregs, *TRGC2^+^ CD8^+^* T cells and *CD4^−^ CD8^−^* T cells were enriched for both CD and UC heritability, whereas *CD8^+^* tissue-resident T cells and gamma-delta T cells were specifically enriched in CD (Fig. 5; FWER < 0.05).

**Fig. 5.**
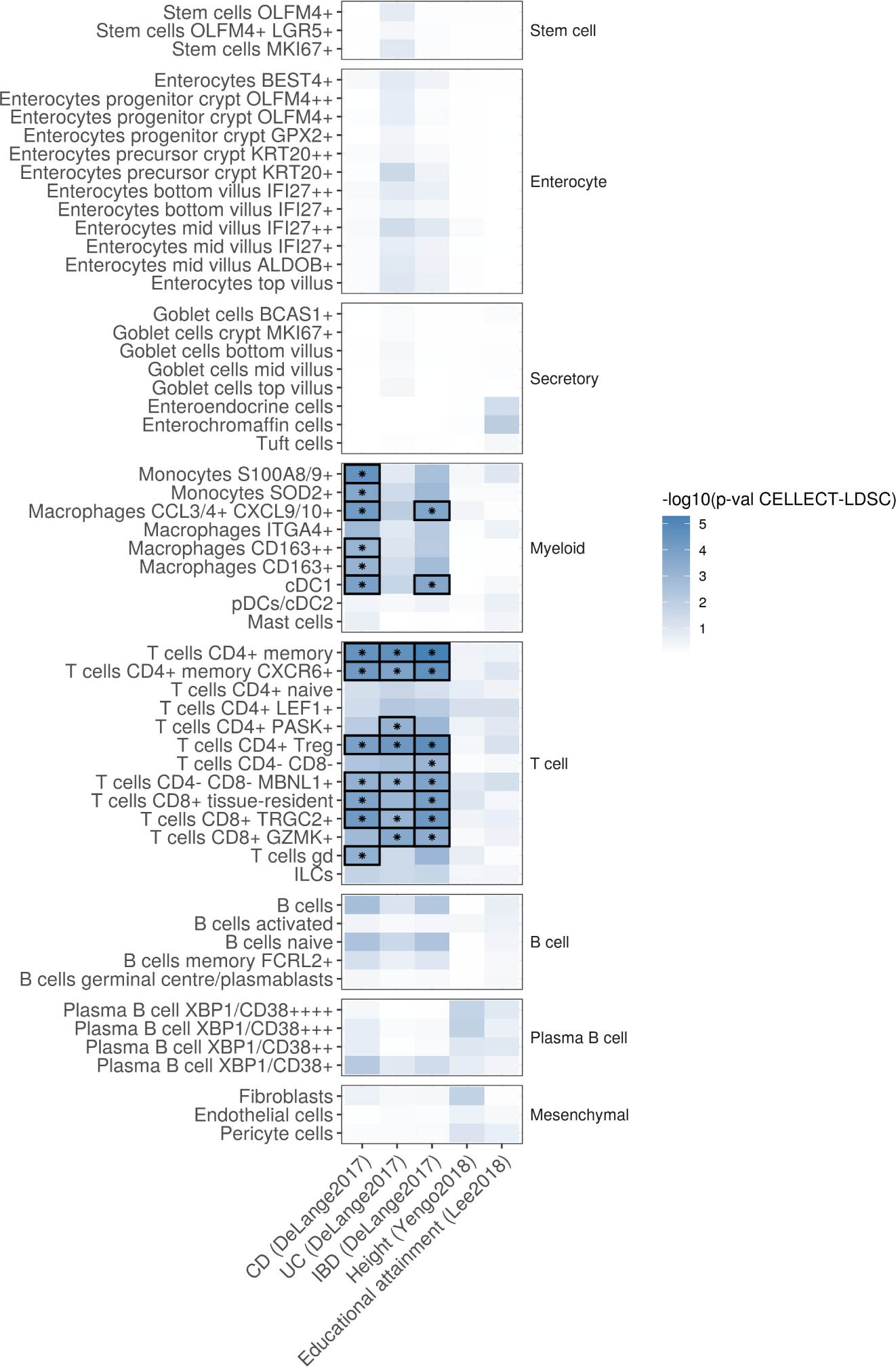
Myeloid and T cells are enriched for CD and UC heritability. Significance of CD, UC and IBD heritability (p-values derived from Stratified LD Score Regression (LDSC)) applied to specifically expressed genes of each cell-type. Results were considered significant at a family-wise error rate FWER < 0.05. Height and educational attainment were used as negative controls.

## Discussion

scRNA-seq is a transformative tool for elucidating the intricate interplay of genes, pathways, and cell types in chronic inflammatory diseases such as Crohn’s disease. However, the inherent biological complexity of these diseases results in significant inter-patient variability in molecular traits like gene expression. This variability is further exacerbated by technical variance—often stemming from poorly understood factors associated with biopsy collection, tissue dissociation, and scRNA-seq protocols—posing substantial challenges to accurately quantifying gene expression differences between groups (e.g., cases vs. controls). These issues are compounded by the historically small sample sizes of scRNA-seq studies and the frequent absence of replication cohorts to validate findings.

To overcome these limitations, we established IBDverse, the largest patient cohort to date for scRNA-seq of terminal ileum biopsies, incorporating both discovery and replication cohorts. We standardized biopsy collection and processing from all 343 patients at a single center, employing consistent protocols for tissue dissociation, single-cell capture, sequencing, and data analysis. Furthermore, we developed a custom auto-annotation model trained on our dataset to reduce technical variability. Despite these rigorous measures, fewer than 50% of differentially expressed genes identified in the discovery cohort were replicated in the validation cohort. Replicability varied markedly across cell types, with poorly annotated or rare populations exhibiting particularly low replication rates. For instance, the scarcity of *S100A8/9*^+^ monocytes and *CXCL9/CXCL10*^+^ macrophages in healthy biopsies limited our ability to reliably detect differentially expressed genes. Larger studies are essential to improve detection of differentially expressed genes in these disease-relevant but underrepresented cell types.

Additionally, the variable transcriptional plasticity of immune cells—shaped by factors such as diet, inflammation severity, disease stage, and treatment regimens—likely contributes to low replication rates. Notably, Crohn’s disease patients in our cohort exhibited wide variation in treatment histories, which likely influenced the observed transcriptomic profiles of immune cells in particular. Our findings underscore the challenges of conducting scRNA-seq differential gene-expression studies on complex, heterogeneous patient populations, and highlight the need for caution particularly when interpreting unreplicated findings from small-scale differential gene expression studies.

We identified a consistent barrier response in the intestinal epithelium, centered on Type I and Type II interferon (IFN) signalling. Genes involved in MHC-I function and Type I IFN pathways were significantly upregulated across the entire crypt-villus axis. This pan-epithelial MHC-I response to barrier damage was largely absent in other cell types and persisted even after resolution of macroscopic inflammation. These findings suggest that inflammation conditions the intestinal barrier, leaving a molecular scar within epithelial progenitors that persists after the resolution of inflammation and likely influences response to future insults. This aligns with recent work by Dennison et al. [51] in pediatric IBD, showing that loss of DNA methylation at MHC-I loci, including NLRC5, enhances MHC-I gene expression in epithelial organoids. We demonstrate that MHC-I expression remains elevated post-inflammation in adult patients, particularly in stem-like progenitor cells such as *OLFM4*^+^ and *MKI67*^+^ populations.

The persistent elevation of MHC-I expression following resolution of macroscopic inflammation is likely to have significant implications for barrier function, perhaps via non-canonical antigen presentation by intestinal epithelial cells (IECs) [52]. Exogenous antigens can access late endosomal compartments in IECs and colocalize with MHC-I proteins in patients with Crohn’s disease (CD), raising the possibility of “cross-presentation.” This process may enable IECs to present luminal antigens to CD8+ T cells, highlighting a potential role for epithelial cells in shaping immune responses. This phenomenon of cross-presentation of exogenous antigen by non-professional APCs has been reported in murine renal epithelium [53], and liver endothelium [54], and in the context of the intestine suggests that epithelial cells play an active role in shaping local inflammatory responses that may be dysregulated in CD.

Epithelial MHC-I expression driven by interferon (IFN) signalling has been observed in various tissues and disease contexts. For instance, during murine *Citrobacter* spp. infection, IFN-γ sensing in the intestinal epithelium facilitates both pathogen and self-antigen presentation to intraepithelial T cells via IRF1 and MHC-I, which suppresses NLRP3 macrophage inflammasome activation. Notably, mice with transgenic deletion of IFN-γ sensing (Ifngr^fl/flVilCre) exhibit reduced IRF1/MHC-I signalling and resistance to anti-TNF therapy [55]. Building on these findings, our data demonstrate that increased epithelial Type II IFN responses are predominantly restricted to enterocytes along the crypt-villus axis and resolve following tissue repair.

Macrophages are among the most abundant leukocytes in the gastrointestinal tract, forming a dense network around the epithelial barrier and playing critical roles in intestinal health [46]. Their dysregulation is strongly associated with chronic inflammatory diseases such as IBD [56], [46]. During active disease, intestinal monocytes and their macrophage progeny accumulate in large numbers, producing pro-inflammatory cytokines like TNF, IL-1β, IL-6, and IL-23 [57], [58], [59]. Although macrophages are hypothesized to be key targets of IBD therapies [46], their heterogeneity, especially in disease states, remains poorly understood. In this study, we identified a transcriptionally distinct population of *ITGA4*^+^ macrophages that become more abundant during intestinal inflammation. Differentially expressed genes in these cells are enriched in nine cytokine pathways, including IL-6, IL-12, and IL-23. This response is underpinned by a core JAK/STAT gene signature, with JAK2 and STAT1/2/3 being significantly differentially expressed. Given that JAK inhibitors, which target multiple cytokine-dependent pathways (e.g., IL-6, IL-10, and IL-23), are now effective therapies for IBD, understanding their cellular targets is critical. Importantly, macrophages from IBD patients carrying disease associated polymorphisms in JAK2 display enhanced JAK2 expression and NOD2-induced JAK2 phosphorylation, and amplified cytokine signalling [60]. Moreover, JAK2-deficient macrophages fail to upregulate MHC-I proteins in response to Type I IFN stimulation, increasing susceptibility to infection and inflammation [61], [62]. We identify *ITGA4*^+^ macrophages as a novel, preferential source of JAK/STAT signalling and downstream cytokine responses in Crohn’s disease, highlighting their potential as therapeutic targets.

Specifically expressed genes from multiple cell-types within the innate and adaptive immune system were enriched with heritability, highlighting their involvement in disease pathogenesis. The strong enrichment of CD heritability among *CXCL9/10^+^* macrophages, *SOD2*^+^ and *S100A8*/*9*^+^ monocytes is particularly noteworthy given both populations are significantly expanded in disease and highly express inflammatory cytokines and known CD susceptibility genes such as *NOD2, PTAFR* and *LRRK2*. Unfortunately, we were unable to detect many reproducible differentially expressed genes in these important cell types, likely due to the rarity of these cells in biopsies from healthy individuals. Whilst we did not identify heritability enrichment within non-immune cell populations, this does not mean that these cells do not contribute to disease pathology. We note that heritability enrichment in non-immune cells has been reported previously, for example within M cells in UC [63]. Our data shows that several likely effector genes within IBD associated loci are specifically expressed by non-immune cells, including *RNF186* [64], *FUT2* [65], *PDLIM5* [4] and *HNF4A* [66]. The underrepresentation of stromal cells in our atlas due to a combination of using pinch biopsies and digestive enzymes that favour epithelial cell capture likely also reduces our power to detect enrichments in these cell types.

In summary, we present IBDverse, the largest single-cell RNA sequencing (scRNA-seq) dataset of terminal ileal biopsies, comprising over 1.1 million cells from 111 Crohn’s disease (CD) patients and 232 healthy controls. Using both discovery and replication cohorts, the study identifies and validates differentially expressed genes, pathways, and cell types associated with CD. We find widespread epithelial changes driven by interferon signalling and persistent MHC-I upregulation post-inflammation, and a distinct population *ITGA4* expressing macrophages that are major contributors to JAK/STAT signalling and cytokine production. Heritability analysis highlights the involvement of immune cell populations, such as *CXCL9/10*^+^ macrophages and *S100A8/9*^+^ monocytes, in CD pathogenesis. These results provide a valuable resource for understanding CD mechanisms and identifying potential therapeutic targets. An open-access portal for navigating our single-cell data resource has been launched at https://www.ibdverse.info/.

## Acknowledgements

This research was supported by the NIHR Cambridge Biomedical Research Centre (BRC-1215-20014). The views expressed are those of the authors and not necessarily those of the NIHR or the Department of Health and Social Care. This research was funded in part by the Wellcome Trust [Grant numbers 206194 and 108413/A/15/D], The Crohn’s Colitis Foundation Genetics Initiative [Grant numbers 612986 and 997266] and Open Targets [OTAR2057].

We thank all individuals who kindly donated samples and their time to the study. We thank Vladimir Kiselev and Martin Prete for setting up the cellxgene interactive data explorer and Henry J Taylor for input on differential gene expression analysis. We also thank Kylie R James for her assistance with providing cell type markers.

## Author contributions

Statistical analysis and manuscript drafting, M.K., T.A., D.L.T. and G.R.J.; Sample collection, K.A., W.G., B.B., and C.Q.C.; Sample processing, M.H.G., M.S., N.W., J.S., J.O., M.X.H., K.A.C., and R.E.M.; Clinical information, N.N.; Critical discussion M.J.P., L.R.N, R.E.M., R.S., V.P., C.P.J., C.C., and M.P.; Data processing M.T., M.O., G.N., S.L., V.I., and Y.G; Writing – Review & Editing, D.C., B.T.H., R.E.M., T.R. and C.A.A.; Conceptualization and Supervision, T.R., C.A.A.

## Materials and Methods

### Sample ascertainment

This study was approved by the National Health Service (NHS) Research Ethics Committee (Cambridge South, REC ID 17/EE/0338). Written informed consent was given by all participants.

Individuals undergoing routine endoscopic assessment were recruited at Addenbrooke’s hospital, Cambridge, UK. Clinical information and metadata for the participants are provided in Table S1. All CD participants classified as ‘inflamed’ had a confirmed history of CD and macroscopic evidence of terminal ileal inflammation from tissue sampled during the biopsy. All control participants were undergoing endoscopic assessment or surveillance for healthy and non-cancer related reasons (e.g., history of iron deficiency anaemia, family history of colorectal cancer). Control participants did not have macroscopic evidence of intestinal inflammation, a personal history of cancer, and were not in receipt of corticosteroids or any other immune modulating therapy. Patients who were taking probiotics or antibiotics were excluded. Patients of non-European ancestries were also excluded to reduce confounding. Pinch-biopsies of the terminal ileum were collected from all participants and deposited into pre-chilled Hanks Balanced Salt Solution (HBSS) without Mg^2+^, Ca^2+^, or phenol red. Samples were placed on ice and immediately transferred to the Sanger Institute.

### Single-cell RNA isolation and sequencing

Terminal ileal biopsies were dissociated using a single-step digestion protocol on ice to release all major intestinal cell types present in the biopsy (epithelial, immune, and stromal) without stressing the cells. First, the biopsies were mechanically minced and pipetted to release immune cells (fraction 1) from the lamina propria and the remaining tissue chunks were transferred to HBSS^−/−^ containing 2 mM EDTA, 0.26 U/µl serine endoprotease isolated from *Bacillus licheniformus* (Sigma, P5380), 5 µM QVD-OPh (Abcam, ab141421), and 50 µM Y-27632 dihydrochloride (Abcam, ab120129). Tissue chunks were pipetted regularly during a 30 minute incubation on ice to release epithelial and stromal cells (fraction 2). The cells from both fractions are washed, centrifuged, and then incubated for 10 minutes at room temperature in Hank’s Balanced Salt Solution (HBSS) with Mg^2+^, Ca^2+^, and without phenol red—including 5 mM CaCl_2_, 1.5U/µl collagenase IV (Worthington, LS004188), and 0.1 mg/ml DNase I (Stem Cell Technologies, 07900). The cells were then filtered (30 µm; CellTrics 04-0042-2316), washed, and centrifuged before being incubated for 3 minutes at room temperature in the red blood cell lysis buffer (ACK lysis buffer; Gibco, A10492). Two final washes and centrifugations were performed before a final filtration (40 µm) and manual cell counting (haemocytometer, NanoEnTek, DHC-N01).

Single-cell RNA sequencing was undertaken using 3’ 10X Genomics kits (v3.0 and v3.1) according to the manufacturer’s instructions. All samples sequenced under kit version v3.1 had dual indexes, samples sequenced under kit version v3.0 had either single or dual indexes. We targeted 6,000 cells for CD participants and 3,000 cells for controls to account for the increased cellular heterogeneity in CD biopsies. Since the proportions of immune cells vary with inflammation status, we altered the ratio of cells from fractions 1 and 2 in an attempt to make the representation of cell types more equal. Viability of the mixed populations was 92±9% (mean ± S.D.) according to Trypan blue staining. Libraries were sequenced using a HiSeq4000 sequencer (Illumina; N_CD_=20, N_control_=4) or NovaSeq S4 XP sequencer (Illumina; N_CD_=91, N_control_=228) with 100bp paired-end reads, targeting 50,000 reads per cell. We compared the fraction of reads mapped confidently to the transcriptome (output metric from CellRanger) within CD and healthy participants and found no difference between sequencers (minimum p-value > 0.05, Wilcoxon rank sum test).

### Single-cell RNA-seq processing and quality control procedures

CellRanger v7.2.0 was used to demultiplex reads, align reads to GRCh38 with Ensembl version 93 transcript definitions (GRCh38-3.0.0 reference file distributed by 10X Genomics), and generate cell by gene count matrices. CellBender v2.1 [67] was then applied to identify droplets containing cells and adjust the raw counts matrix for background ambient transcript contamination. For training, CellBender requires a rough estimate of the number of droplets containing cells (cell droplets) and the number of droplets without cells (empty droplets) derived from the UMI curve—the rank ordering droplet barcodes according to total UMI counts (x axis) by the total number of UMI counts per droplet (y axis). The UMI curve was calculated from droplets with a UMI count >1,000, and the threshold estimated using the “barcoderanks-inflection” procedure from DropletUtils v1.9.16 [68]. To estimate the number of empty droplets, we calculated the UMI curve as described above, selected droplets with a UMI count between 250 and 10, and estimated the threshold by performing both the “barcoderanks-inflection” and “barcoderanks-knee” procedure from DropletUtils—using 1/3rd of the distance between the two estimates as the final threshold. CellBender was run with default parameters except for excluding droplets with <10 UMI counts (--low-count-threshold) and using 300 epochs with a learning rate of 1×10^−7^. The final counts matrix was adjusted for the ambient transcript signature at a false positive rate of 0.1. Next, multiplets were identified and removed using scrublet v0.2.1 [69], simulating 100,000 multiplets and calculating the multiplet threshold using the threshold_li function from the scikit-image package v0.17.2 [70], initialised using the threshold_otsu function. The reported sex of each sample was verified by generating pseudobulk expression matrices and comparing the expression of *XIST* to the mean expression of all genes on the Y chromosome.

### *De novo* cell type identification

The atlasing cohort was used to identify cell types and fit a model to automatically predict cell types across the entire dataset. First, additional filters were applied to ensure only the highest quality cells were used for *de novo* clustering. Cells with fewer than 100 genes expressed at ≥ 1 count, or where the percentage of counts originating from the mitochondrial genome (https://www.genenames.org/data/genegroup/#!/group/1972) was > 50, were removed. Next, an isolation forest (scikit-learn v0.23.2) was used to remove outlier cells based on (i) the percentage of counts originating from the mitochondrial genome, (ii) the total number of UMI counts per cell, (iii) the number of genes expressed (>=1 count) per cell. These metrics were selected following the recommendations in [71].

Subsequent processing and management of the expression data was performed using scanpy v1.6.0 [72]. Genes expressed (>=1 count) in five or fewer cells across the whole dataset were removed (sc.pp.filter_genes with min_cells=5). To account for variable sequencing depth across cells, unique molecular identifier (UMI) counts were normalised by the total number of counts per cell, scaled to counts per 10,000 (CP10K; sc.pp.normalise_per_cell), and the CP10K expression matrix (ln[CP10K+1]; sc.pp.log1p) was log-transformed.

To perform dimensionality reduction, the 2,000 most variable genes across samples were selected by (i) calculating the most variable genes per sample and (ii) selecting the 2,000 genes that occurred most often across samples (sc.pp.highly_variable_genes with flavor=’seurat’ and batch_key=sample). After mean centering and scaling the ln(CP10K+1) expression matrix to unit variance, principal component analysis (PCA; sc.tl.pca) was undertaken using the 2,000 most variable genes after removal of protein coding mitochondrial, ribosomal, and immunoglobulin genes, because these genes constituted the ambient signature learned by CellBender. To select the number of PCs for subsequent analyses, we used a scree plot [73] and calculated the “knee/elbow” derived from the variance explained by each PC using the kneedle estimator v0.7.0 [74]. From the automatically estimated elbow, we included five additional PCs in order to ensure all meaningful variability was captured, selecting 29 PCs for clustering. Finally, bbknn v1.3.12 [75] was applied to integrate samples and control for sample specific batch effects.

Clusters were defined using the Leiden graph-based clustering algorithm v0.8.3 [76] on the nearest neighbours determined by bbknn. Clusters were generated across a range of resolutions from 0.5 to 5 to empirically determine the optimal clustering resolution. For each resolution considered, the data was divided into training (2/3 of cells) and test (1/3 of cells) sets and a single layer dense neural network fit to predict cluster identity from expression using keras v2.4.3. The cluster label of each cell was predicted and the Matthews correlation coefficient (MCC) calculated for each cluster [77]. The final cluster classifications were chosen to achieve a minimum MCC of > 0.75 across all clusters, with a resolution of 3.25 selected to meet this criterion (Fig. S9). At this resolution, all clusters met the threshold except for cluster 40, which exhibited MCC < 0.75 at many resolutions so was excluded (Fig. S9). This adjustment yielded a total of 57 clusters.

### Cell type annotation

To determine the cell type identity of the 57 clusters, marker genes for each cluster were identified using the Wilcoxon rank-sum test (sc.tl.rank_genes_groups with method=’wilcoxon’) to compare the gene expression of each cell type to all other cell types and rank genes according to differences in expression. Highly discriminative marker genes with a Bonferroni-corrected p-value <0.05 were then used to label cell types through expert knowledge. To further visualise the annotated cell types, dimensionality reduction was undertaken using the uniform manifold approximation and projection (UMAP) algorithm, implemented within scanpy (scanpy.tl.umap) with default parameters, except for changing the minimum distance from 0.5 to 1.0. Analysing all cells that passed QC, we identified eight major cell populations including epithelial cells (stem cells, enterocytes and secretory cells), immune cells (T and B cells, plasma B cells and Myeloid cells), and mesenchymal cells.

Within epithelial cells, we identified three distinct stem cell populations: *OLFM4^+^* stem cells, *OLFM4^+^ LGR5^+^*stem cells, and proliferating *MKI67^+^* stem cells. Among enterocytes, we identified progenitor cells marked by *OLFM4* and *GPX2*, precursor cells expressing *KRT20*, and a range of enterocytes expressing *IFI27*. We further distinguished enterocytes along the crypt-villus axis (crypt, middle, top) based on signature genes such as *ALPI, APOA4,* and *APOC3* [22]. In the secretory cell lineage, we identified goblet cells (*CLCA1, FCGBP, MUC2*), including proliferating *MKI67^+^* and *BCAS1^+^* goblet cells, as well as goblet cells positioned along the crypt-villus axis, marked by *EGFR, KLF4, NT5E*, and *SLC17A5*. Additional cell types included enteroendocrine cells (*NTS, PYY, GCG*), enterochromaffin cells (*TPH1, CES1*), and tuft cells (*PLCG2, PTGS1, LRMP*). A summary of these markers and their expression across cell types is visualised in Fig. S1.

Within immune cells we identified monocytes expressing *S100A8/9, SOD2, and CXCL9/10,* macrophages with positive expression of ITGA4, resident macrophage populations (*CD163, MAF, C1QA/B/C*), conventional dendritic cells type 1 cDC1 (*XCR1, BATF3*), a mix of plasmacytoid dendritic cells and conventional dendritic cells type 2 (pDC/cDC2) determined by (*IRF4, ZEB1, FLT3*) and mast cells (*MS4A2, TPSAB1*) (Fig. S1). Additionally, we identified thirteen distinct T cell populations, including *CD4^+^* and *CD8^+^*T cells, innate lymphoid cells (ILCs, marked by *IL1R1, ALDOC, LSTI*), and gamma-delta T cells (*TRGC1, TRDC, GZMA/B*). Within the *CD4^+^* T cell subset, we characterized naive T cells (*SELL, CCR7*), *LEF1^+^* and *PASK^+^* expressing *CD4^+^* T cells, regulatory T cells (Tregs, marked by *FOXP3, TIGIT*), and two populations of double-negative (*CD4^−^ CD8^−^*) T cells, one of which shows elevated *MBLN1* expression. In the *CD8^+^* T cell subset, we identified two populations of tissue-resident *CD8^+^* T cells expressing *TRGC2,* as well as a distinct population of GZMK*^+^* expressing *CD8^+^* T cells (Fig. S1).

Among B cells, we identified *FAU* expressing B cells, activated B cells (*CKS1B, STMN1*), naive B cells (*IGHD/M, FCER2*), and germinal center/plasmablast B cells (*CD19, CD38, TCL1A*). Additionally, we observed a gradient of plasma cells with varying levels of *XBP1* and *CD38* expression (Fig. S1).

Lastly, we identified three mesenchymal cell populations: fibroblasts (*COL1A1/2, COL3A1*), endothelial cells (*PECAM1, VWF*) expressing *ACKR1*, and pericytes (*PDGFRB, CSPG4*) (Fig. S1).

We did not detect a distinct group of neutrophil and eosinophil cells - as previously well documented, they are poorly represented due to the limited ability of the 10X scRNA-seq process to capture granulocytes [78].

### Crypt-villus score

We positioned epithelial cells along the crypt-villus axis, identifying top-villus epithelial cells as well as enterocyte precursors/progenitors and goblet cells located at the crypt base. Stem cells and enterocytes were scored based on expression of genes such as *APOA4, APOC3, ALPI, PKIB, PMP22,* and *SLC28A2*, derived from spatial transcriptomics data that characterize crypt and villus intestinal cells [22]. Similarly, we applied signature genes *EGFR, KLF4, NT5E,* and *SLC17A5* to score secretory cells across the crypt-villus axis.

### Automatic cell type annotation

To annotate cell types across all samples, we used Celltypist v1.6.2 [79] to train a classification model based on our identified clusters. This trained Celltypist model was then applied to assign cell type labels to all 343 scRNA-seq samples, following the initial processing steps detailed in the “Single-cell RNA-seq processing and quality control” section. Cells with a Celltypist confidence score below 0.5 were excluded from subsequent analyses.

### Differential gene expression analysis

For each cell type, we tested for association of gene expression with CD disease status or inflammation severity (TI-SES-CD score) using MAST v1.14.0, a two-part, generalised linear model with a logistic regression component for the discrete process (i.e., a gene is expressed or not) and linear regression component for the continuous process (i.e., the expression level) [80]. For gene *i*, individual *j*, and cell *k*, let *Z_ki_* indicate whether gene *i* is expressed in cell *k* and *Y_ki_* denote the ln(CP10K+1) normalised gene expression. A two-part regression model was used to test for association:

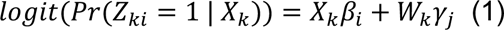

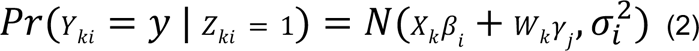

where *X_k_* are the predictor variables for cell *k*, *W_k_* is the random effect design matrix of cell *k* belonging to individual *j*, *β_i_* is the vector of fixed effect regression coefficients, and *γ_j_* is the vector of random effects (i.e., the random complement to *β_i_*), normally distributed with mean zero and variance *σ*^2^_*γk*_. In all DE comparisons, whether contrasting CD inflamed or uninflamed with healthy controls or examining association of gene expression to TI-SES-CD score - sex, age (binned into groups of five), cell mitochondrial percentage (technical covariate associated with cellular stress), and cell complexity i.e., the number of genes detected per cell [80], [14] were included as fixed effect variables and individual as a random effect to control for pseudoreplication bias [81]. The Benjamini-Hochberg procedure [82] was used to control for multiple testing across all cell types, and p-values were obtained from the hurdle model, derived from the summed *χ^2^* null distributions of the discrete (*Z_i_*) and continuous (*Y_i_*) components, as described in [80]. To increase the speed of each test, genes with an average CP10K of <1 in that cell type were removed prior to fitting models for each cell type.

### Gene set enrichment analysis

Gene set enrichment analyses were performed using GSEA v1.17.1 [83] with default parameters to identify pathways enriched among differentially expressed genes. Pathways were obtained from the reactome v76 gene pathway database [84] as part of the molecular signatures database (MSigDB) v7.4 [85]. Z-scores from the CD vs control differential gene expression hurdle model, were used as input for enrichment analyses.

### Prioritisation of IBD effector genes

Forty four genes likely to be perturbed in IBD were identified from within IBD-associated loci based on a several criteria, including but not limited to 1) presence of a coding mutation fine-mapped down to single variant resolution, 2) detailed and convincing functional follow-up work that established the causality of the gene or 3) the protein encoded by the gene plays a major role in a pathway that is targeted by an existing IBD therapy. Note, it is typically not straightforward to identify disease effector genes from within GWAS loci and this challenge remains a major focus for the field of complex disease genetics. While our list of 45 likely IBD effector genes is undoubtedly greatly enriched for true IBD effector genes, false-positives could still remain.

### Specifically expressed genes

We identified specifically expressed genes (SEGs) for each cell type using CELLEX v1.2.1 package [48]. CELLEX calculates specifically expressed gene scores using four complementary approaches that include Gene Enrichment Score [86], Expression Proportion [87], Normalized Specificity Index [88] and Differential Expression T-statistic - the package produces a normalised mean of these four metrics which we used as our Specificity score.

### Heritability analysis

Heritability enrichment analysis was performed using the CELLECT v1.3.0 workflow [48]. This workflow deploys stratified LD score regression (S-LDSC) [7] to identify cell types with features that are enriched in genetic associations for a disease/trait of interest. CELLECT was run with default parameters, which includes filtering out complex genetic regions such as the HLA locus prior to analysis. CELLECT requires summary statistics from a genetic association study and a set of gene scores (ranging between 0 and 1) for each gene that are to be tested for heritability enrichment. For genetic summary statistics of interest, we used CD and UC statistics from [3] and as negative controls, we used genetic summary statistics for height [49] and [50]. For the gene scores, we used cell type specific gene scores with Bonferroni correction to control for the total number of tests across all cell types.

### Data availability

Raw sequencing data files are available at the European Genome-phenome Archive (https://ega-archive.org), accession number: *Available after publication*. Processed data are available at zenodo (https://zenodo.org), accession number DOI: https://zenodo.org/records/14276773 and through https://www.ibdverse.info/.

### Code availability

The code used for analyses within this study is available at GitHub repository (*Available after publication)*.

## Supplementary Figures and Tables

Fig. S1. Marker gene expression used to curate annotations within the terminal ileum atlas.

Fig. S2. Epithelial cell types represent the crypt-villus axis differentiation.

Fig. S3. Cell-type proportions across healthy and CD samples in the atlasing cohort.

Fig. S4. Concordance of gene specificities across discovery and replication datasets.

Fig. S5. Accuracy in re-annotating the atlas cohort.

Fig. S6. Differentially expressed genes between CD inflamed and healthy samples across all 57 cell types.

Fig. S7. Dysregulated pathways in CD versus healthy epithelial cells.

Fig. S8. Myeloid cell types enriched for CD heritability are found predominantly in CD gut biopsies.

Fig. S9. Optimisation of cluster resolution for cell-type identification.

Table S1. Clinical information for the IBDverse samples.

Table S2. Manually curated list of marker genes for cell types in the atlasing cohort dataset.

Table S3. Demographics of healthy and disease samples across cohorts.

Table S4. Significantly differentially expressed genes within each of the identified 57 cell types in the auto-annotated discovery, replication and full cohort datasets.

Table S5. Correlation between gene-expression fold changes from differential gene expression tests between our discovery and replication cohorts.

Table S6. Dysregulated pathways in CD inflamed epithelial cells.

