## Supplementary figures and tables for "Single-Cell RNA Sequencing of Terminal Ileal Biopsies Identifies Signatures of Crohn’s Disease Pathogenesis"

#### Fig. S1. Marker gene expression used to curate annotations within the terminal ileum atlas.

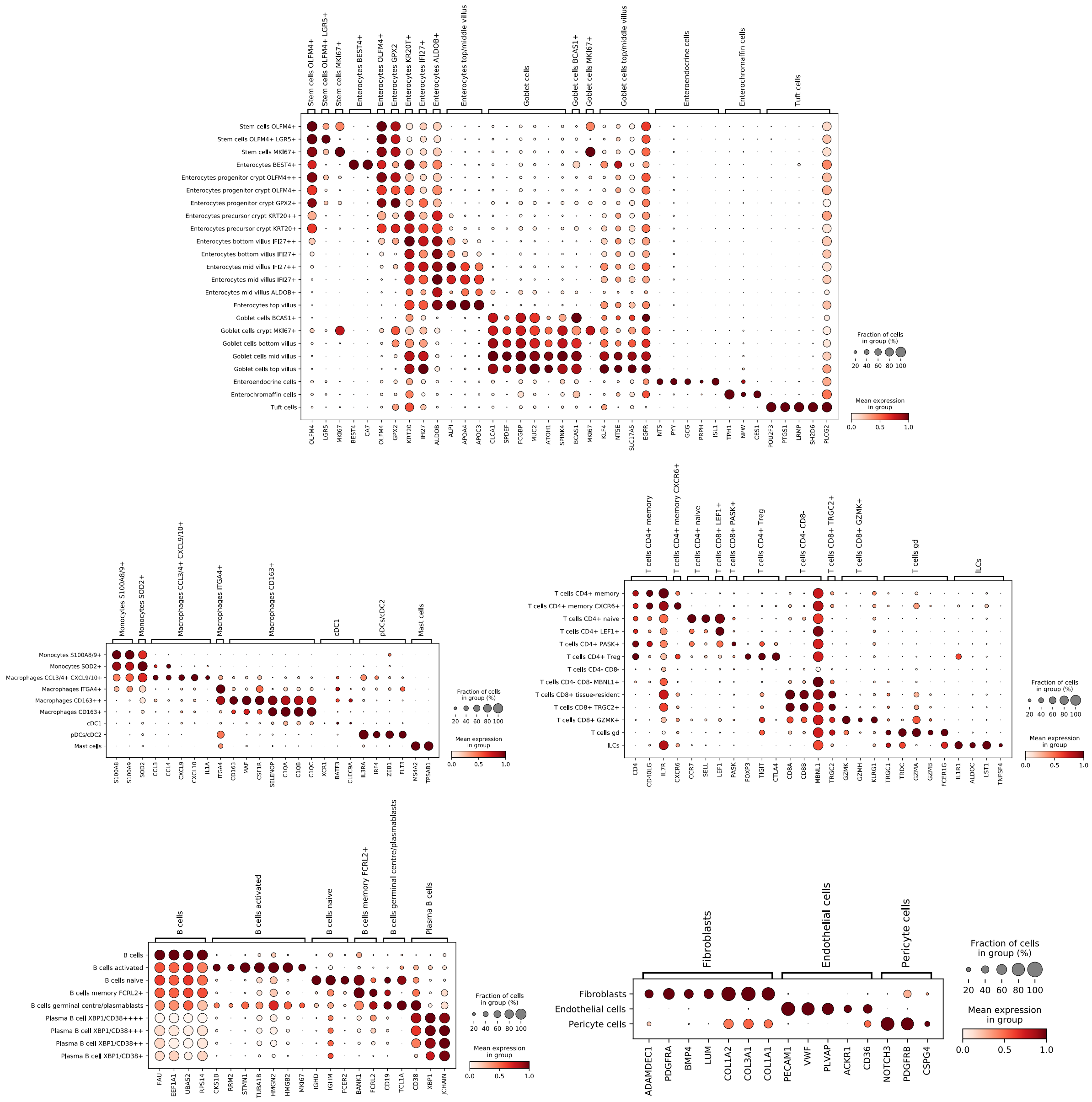


Transcriptional signatures based on literature-derived or expert-curated markers were used to define epithelial, myeloid, T cell, B cell, plasma B cell, and mesenchymal cell types. Dot size indicates the proportion of cells within each cluster expressing a specific marker, color intensity reflects the mean expression of the gene within the cluster.

**Fig. S2. Epithelial cell types represent the crypt-villus axis differentiation.**

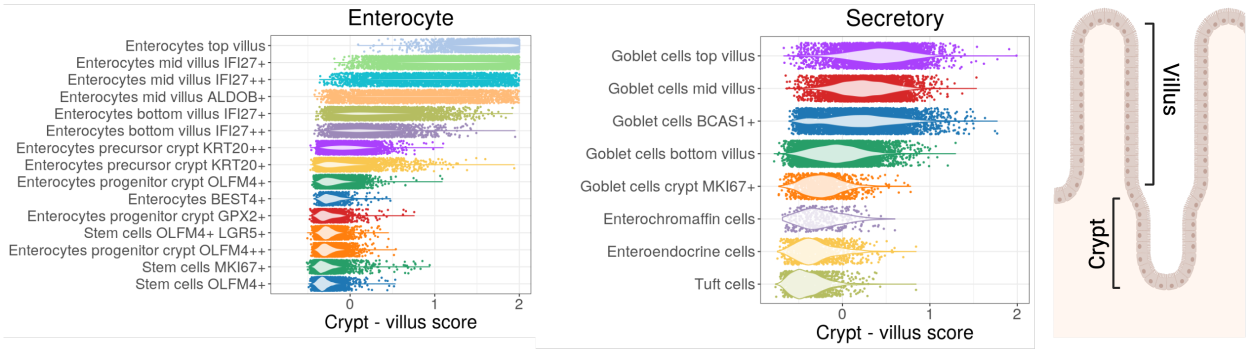


Spatial distribution of enterocytes and secretory cells along the crypt-villus axis was inferred using gene signatures from Moor et al. (2018) (Methods). A subset of enterocytes, termed "top villus enterocytes," exhibited elevated expression scores for a specific gene signature (including *APOA4, APOC3, ALPI*), indicative of cells located at the villus tip. In contrast, enterocytes situated along the mid-villus or in progenitor/stem cell zones showed lower expression of these markers. Similarly, goblet cells were evaluated for a top-villus gene signature comprising *EGFR, KLF4 and NT5E*. Top villus goblet cells demonstrated slightly higher expression scores compared to goblet cells in mid-villus regions and those at the crypt base.

### Fig. S3. Cell-type proportions across healthy and CD samples in the atlasing cohort.

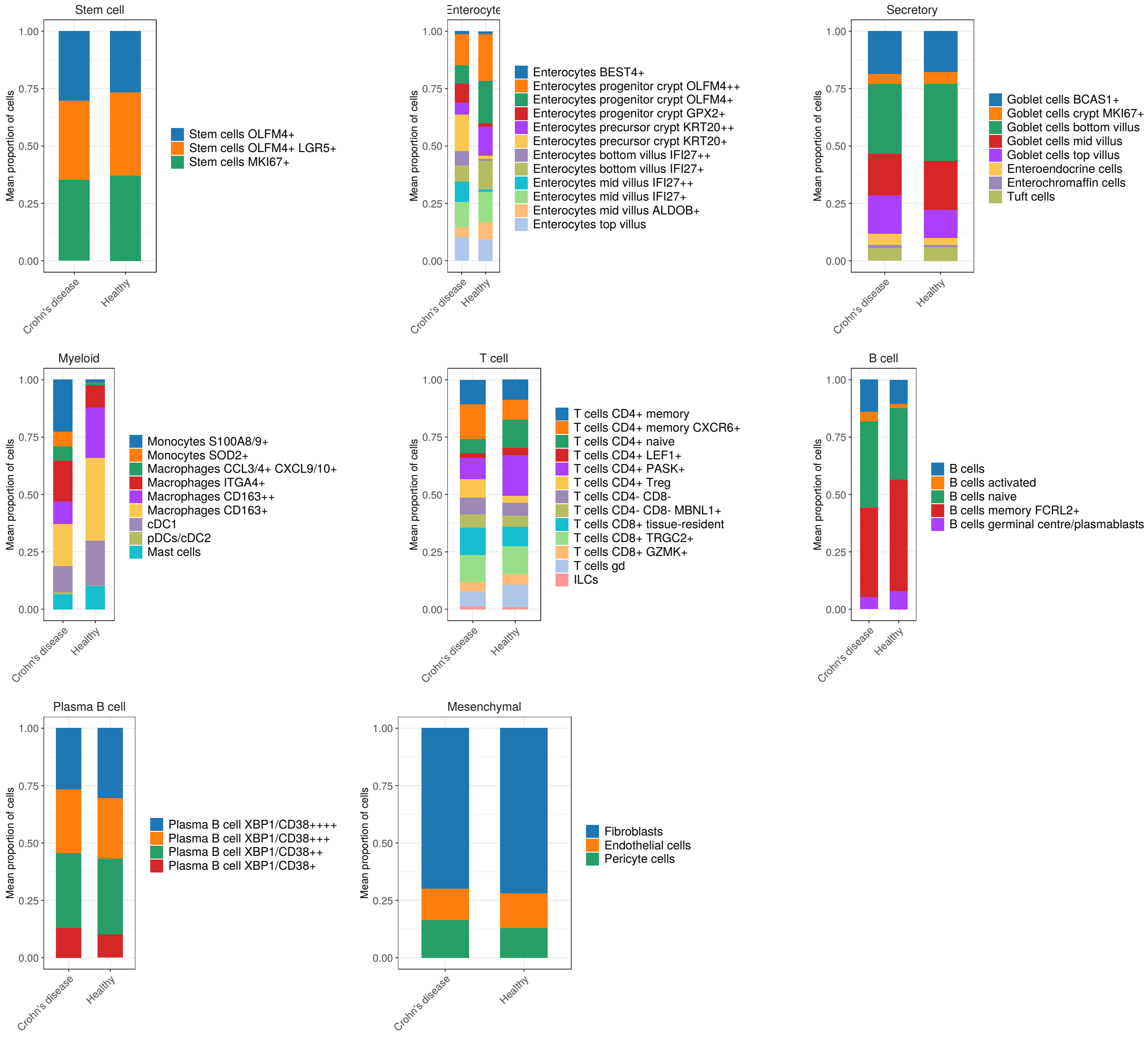


Compositional differences in cell-type proportions between CD patients (n=25) and healthy controls (n=35) across eight major cell populations.

**Fig. S4. Concordance of gene specificities across discovery and replication datasets.**

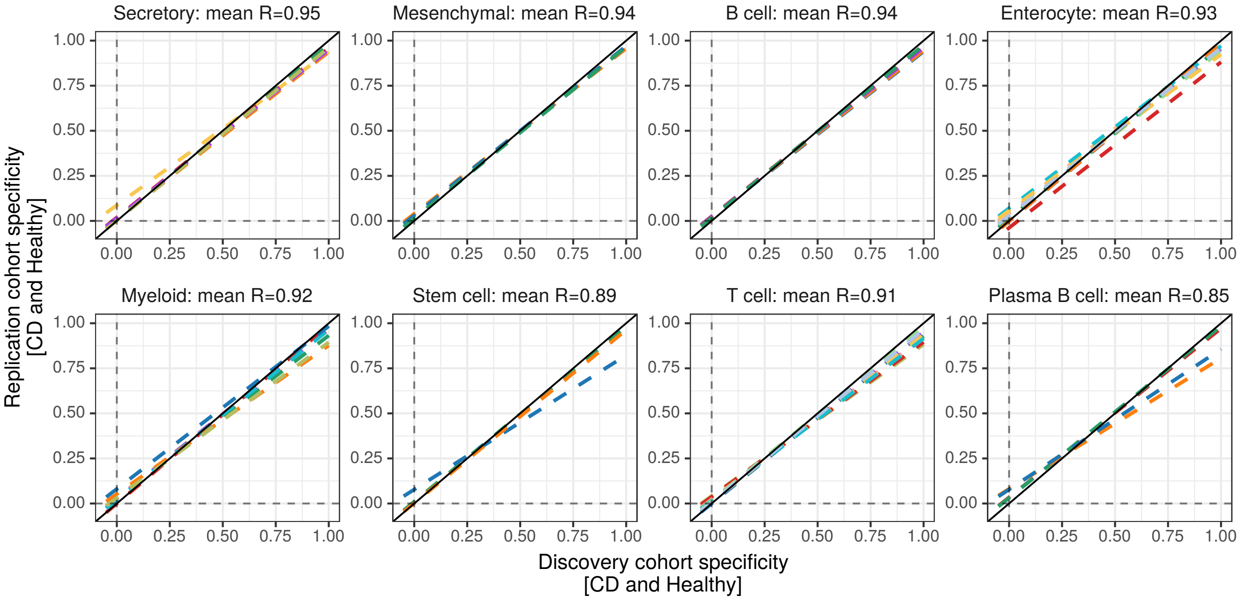


Linear regression model between computationally determined, specifically expressed genes (without thresholding, as outlined in Methods) in the discovery dataset (n=171, x-axis) and replication dataset (n=172, y-axis). For the color legend, please refer to Fig. 1b. The reported mean R represents the average of regression coefficients calculated across cell types within each major cell population.

#### Fig. S5. Accuracy in re-annotating the atlas cohort.

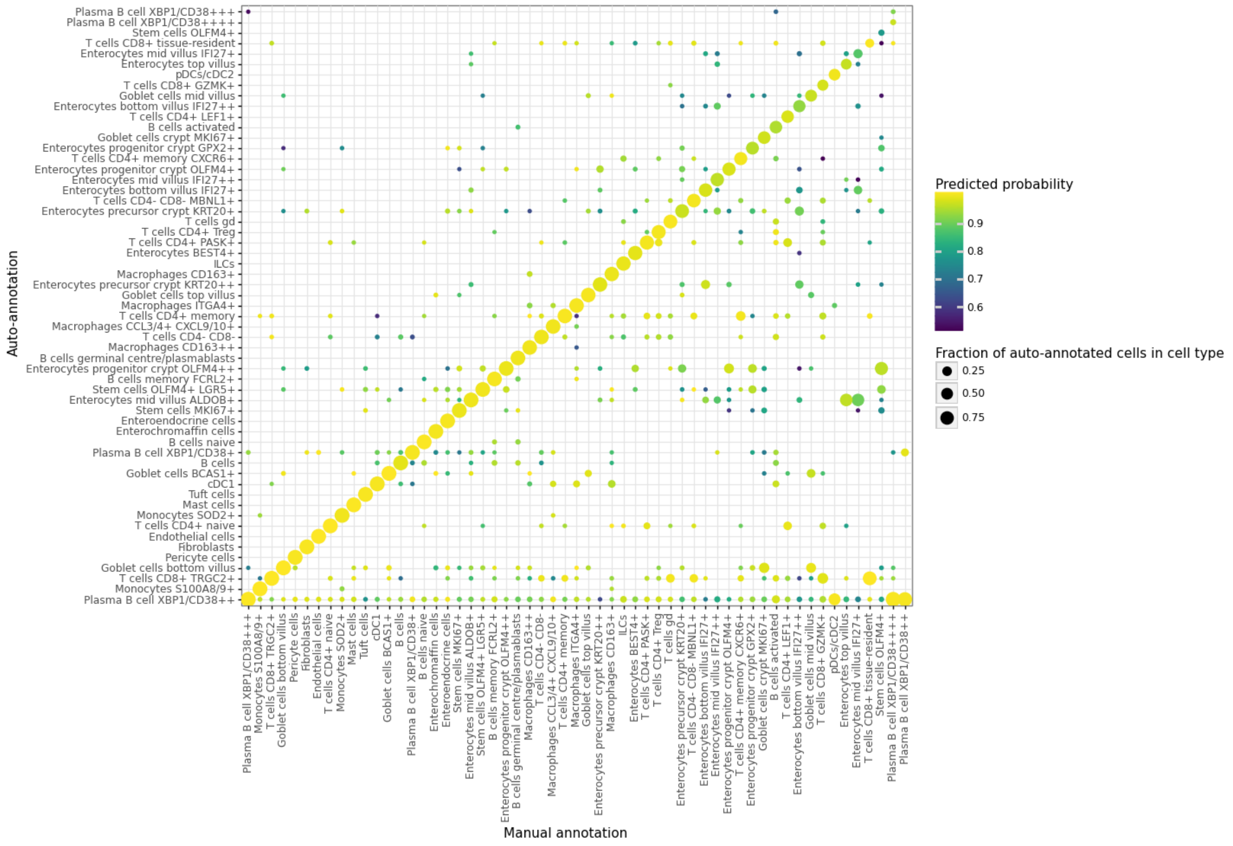


Dot size represents the proportion of auto-annotated cells in the atlas cohort (n=70, y-axis) within each manually defined cluster (x-axis), with color indicating the probability of the mapping prediction.

#### Fig. S6. Differentially expressed genes between CD inflamed and healthy samples across all 57 cell types.

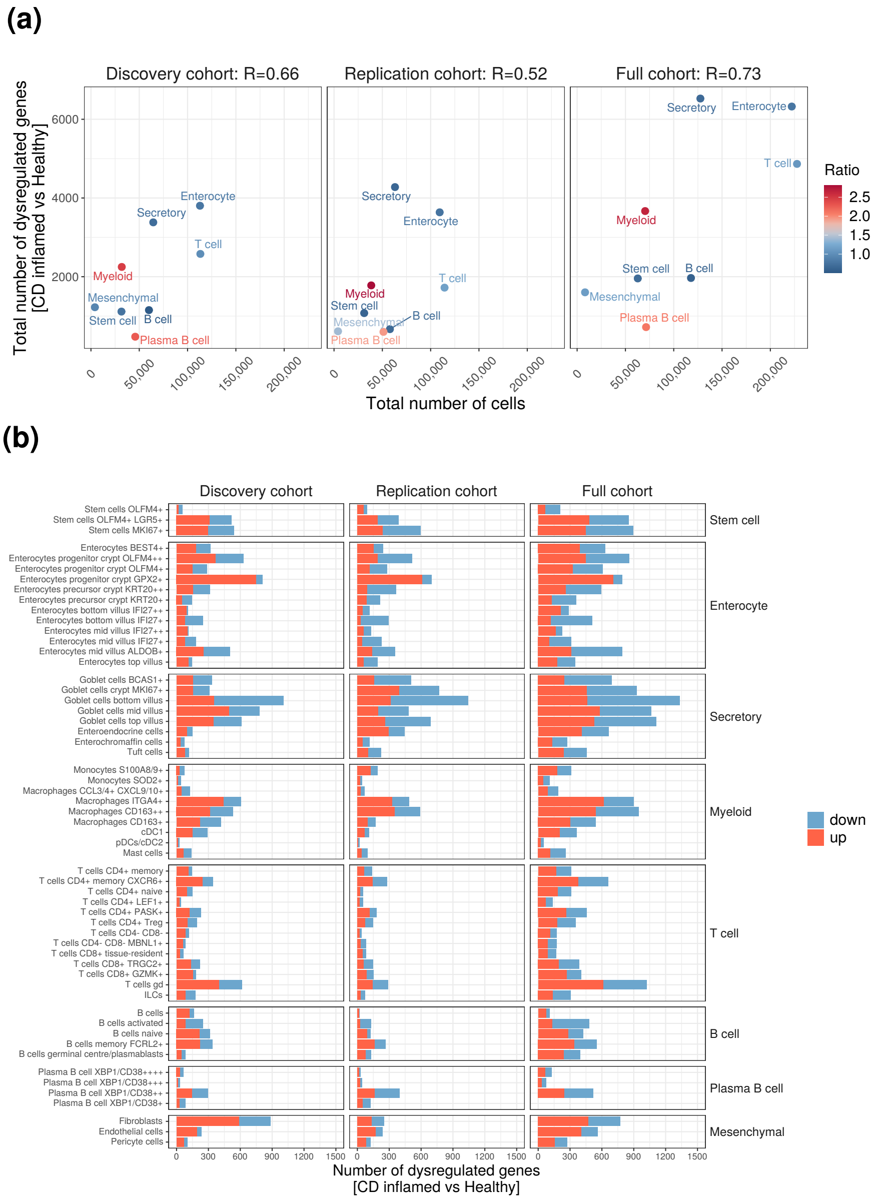


**(a)** The total number of cells in each major cell population (x-axis), the total number of significantly dysregulated genes (FDR $<$ 5%), and the ratio of cells in CD versus healthy controls (color-coded) across the discovery, replication, and full cohorts. Pearson correlation coefficients (R) were calculated for all major cell populations. **(b)** The number of significantly up- and down-regulated genes (FDR $<$ 5%) is shown on the x-axis for each of the 57 cell types (y-axis) across the discovery, replication, and full cohorts.

#### Fig. S7. Dysregulated pathways in CD versus healthy epithelial cells.

**(a)** MHC-I antigen presentation

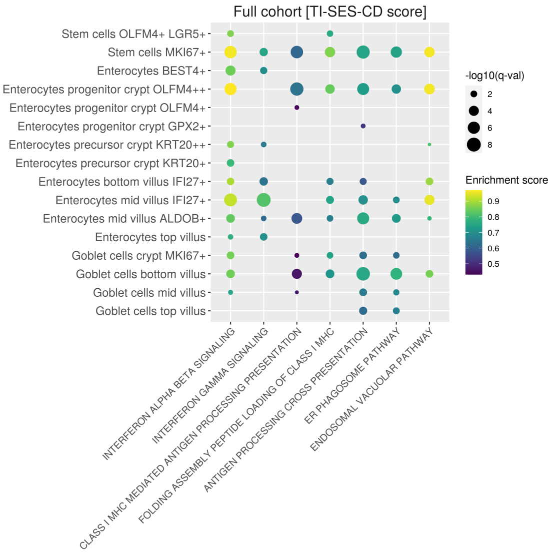


**(b)** MHC-I vs MHC-II antigen presentation

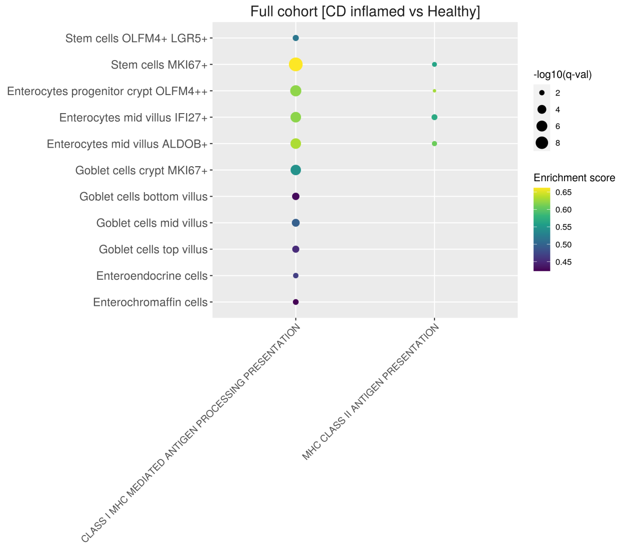


**(c)** Metabolic pathways

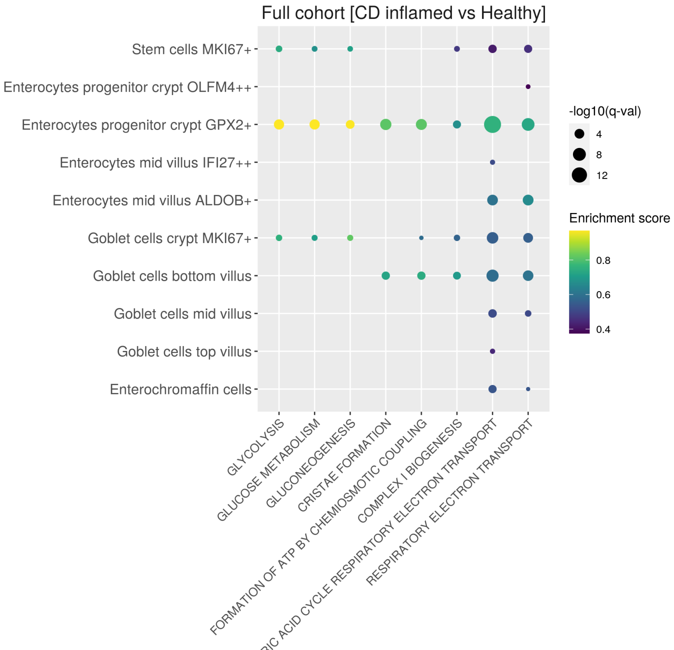


Gene set enrichment analysis was conducted on z-scores derived from the differential gene expression hurdle model applied to the full cohort. The results reveal upregulated pathways in CD epithelial cells, including **(a)** those associated with the TI-SES-CD score, **(b)** enrichment of MHC class II antigen presentation compared to MHC class I, and **(c)** significant metabolic changes within epithelial cells. Populations exhibiting low fold-change replicability ($R < 0.5$, Table S5) were excluded from this analysis.

**Fig. S8. Myeloid cell types enriched for CD heritability are found predominantly in CD gut biopsies.**

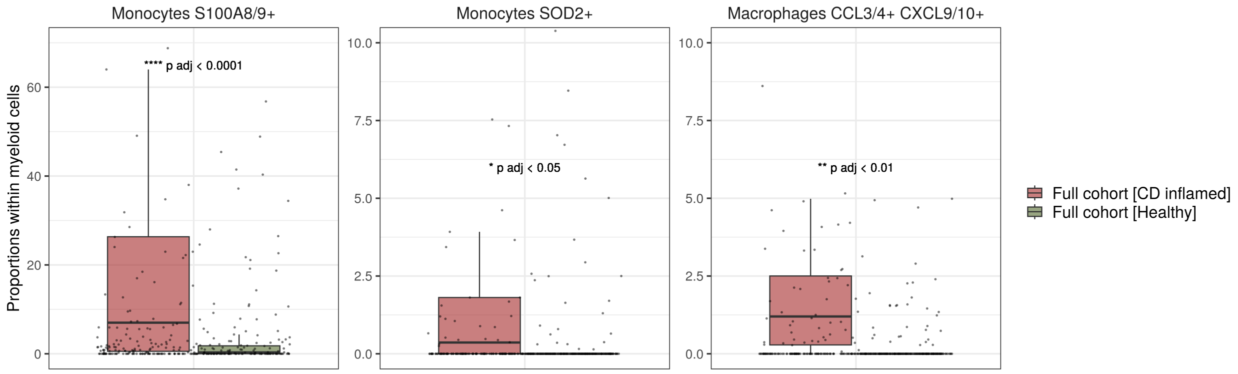


Boxplots show statistical significance (t-test) in the proportions of *S100A8/9+* and *SOD2+* monocytes and *CCL3/4+ CXCL9/10+* macrophages within myeloid cells, comparing inflamed CD and healthy samples.

#### Fig. S9. Optimisation of cluster resolution for cell-type identification.

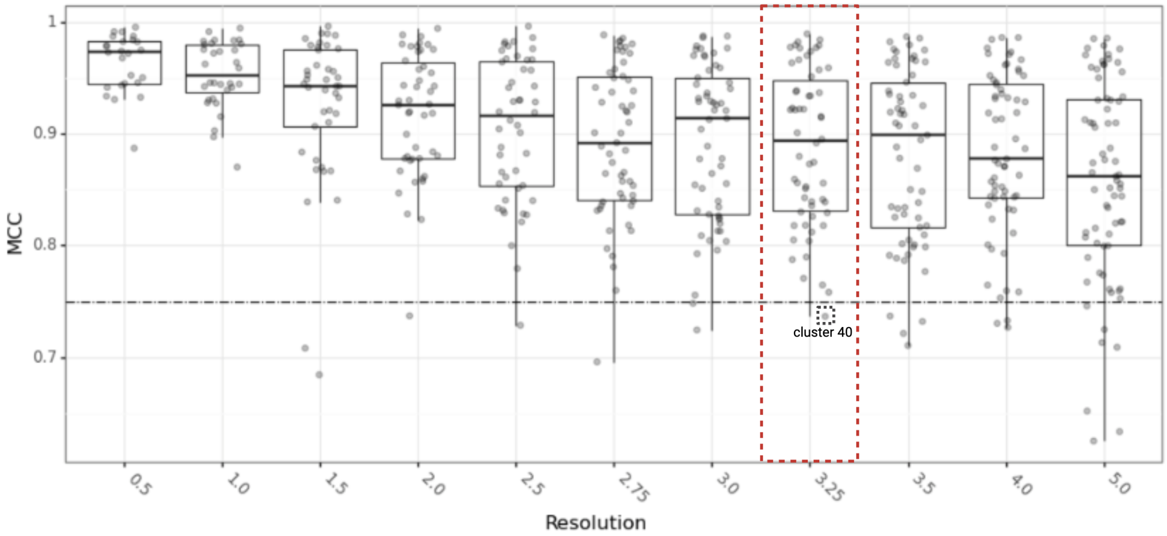


Cluster predictability (x-axis) across a range of clustering resolutions (y-axis). Cluster predictability was assessed by training a keras model on 2/3 of data and calculating the matthews correlation coefficient (MCC) in the remaining 1/3 test set (Methods). A resolution of 3.25 was selected, as cluster predictability rapidly declined at resolutions greater than 3.25. At this resolution, all clusters met the MCC threshold of $> 0.75$, with the exception of cluster 40, which exhibited an MCC $< 0.75$ at multiple resolutions and was therefore excluded.

### Supplementary Tables

**Table S3. Demographics of healthy and disease samples across cohorts.**

| | Discovery (%) | Replication (%) | P-value |
|---|---|---|---|
| N | 171 | 172 | |
| Disease status = Crohn's Disease | 57 (33.3) | 54 (31.4) | 0.730 |
| Inflamed | 29 (17.0) | 35 (20.3) | 0.489 |
| Sex = M | 82 (48.0) | 84 (48.8) | 0.914 |
| Mean age (SD) | 46.62 (14.05) | 46.49 (12.98) | 0.928 |

Absolute number and proportions of demographics across discovery and replication cohorts. Non-significant differences determined by Fisher's exact test or t-test (for age). SD=standard deviation.
